# Patient stratification reveals the molecular basis of disease co-occurrences

**DOI:** 10.1101/2021.07.22.21260979

**Authors:** Beatriz Urda-García, Jon Sánchez-Valle, Rosalba Lepore, Alfonso Valencia

**Author notes:** Alfonso Valencia and Beatriz Urda-García., **Email:**. **Competing Interest Statement:** None declared.

## Abstract

Epidemiological evidence shows that some diseases tend to co-occur; more exactly, certain groups of patients with a given disease are at a higher risk of developing a specific secondary condition. Here we develop a new approach to generate a disease network that uses the accumulating RNA-seq data on human diseases to significantly match an unprecedented proportion of known comorbidities, providing plausible biological models for such co-occurrences and effectively mirroring the underlying structure of complex disease relationships. Furthermore, 64% of the known disease pairs can be explained by analyzing groups of patients with similar expression profiles, highlighting the importance of patient stratification in the study of comorbidities. These results solidly support the existence of molecular mechanisms behind many of the known comorbidities, with most captured co-occurrences implicating the immune system. Additionally, we identified new and potentially underdiagnosed comorbidities, providing molecular insights that could inform targeted therapeutic strategies. We provide a functional and comprehensive resource to explore diseases, disease co-occurrences and their underlying molecular processes at different resolution levels at http://disease-perception.bsc.es/rgenexcom/.

**Significance Statement:** Understanding why certain diseases tend to co-occur is key for improving patient outcomes. This study introduces a new approach to map disease co-occurrences using large-scale RNA sequencing data, revealing that many comorbidities share a molecular basis, often linked to the immune system. By stratifying patients based on similar gene expression profiles, we uncover additional known disease associations and suggest potential new ones, highlighting the need for personalized approaches to comorbidities. These findings, accessible through a web resource, provide a framework for systematically exploring diseases and their relationships at the molecular level, potentially leading to more effective management and treatment strategies.

## Introduction

Disease comorbidity is defined as the co-occurrence of two or more conditions in the same patient (1). Comorbidity incidence increases with age and has a high impact on life expectancy, as it increases patient mortality and complicates the choice of therapies, posing a major problem for patients and health care systems. Accumulating evidence from epidemiological studies indicates that such co-occurrences do not appear randomly and that specific trends are observed, with some diseases co-occurring more than expected by chance (2, 3). Systematic studies using electronic health records have been performed to analyze comorbidity patterns in a given population where disease co-occurrences are represented by static networks (2, 3) or network trajectories if their progression over time is considered (4). These studies demonstrated the predictive value of comorbidity patterns to determine disease progression and outcome, including mortality risk. Crucially, they also showed that patients suffering from a given disease often present different risks of developing specific secondary conditions, underscoring the need for a deeper understanding of how diseases interact and lead to others (4).

The observed patterns suggest that co-occurring diseases might share underlying molecular mechanisms and risk factors, which can be both genetic and environmental, such as drug exposure and lifestyle. Thus, a better understanding of the molecular mechanisms behind comorbidities is a crucial step towards improved prevention, diagnosis, and treatment of these conditions.

Recent studies on comorbidities have included molecular information often analyzing pairs of diseases based on shared disease-related genes (5). Similar to functionally related genes, disease-associated genes tend to colocalize in the protein-protein interaction network forming disease modules which can aid the identification of novel candidate genes and inform about disease associations, including phenotypic similarity and comorbidities (6). In this context, previous work showed that the overlapping gene expression signatures between several Central Nervous System disorders (CNSd) and cancer types could inform about the molecular mechanisms underlying their direct and inverse comorbidities (7, 8). More recently, we have observed that disease similarity networks based on gene expression profiles can be used to identify known comorbidity relationships (8). Although these efforts were able to capture interesting examples, they were unable to recapitulate a large proportion of what is known at the medical level. The mentioned approaches based on PPIs and microarrays reproduced a very small percentage of the epidemiology, and other networks based on miRNAs (9), the microbiome (10) or subcellular localization (11) had no capacity for it. Hence, we address the still largely unknown extent to which molecular information can provide a general explanation to comorbidities.

Here, we reformulate the problem and show that gene expression data is able to reproduce a substantial percentage of medically known disease co-occurrences. Leveraging publicly available RNA-seq data sets, which offer enhanced sensitivity, reproducibility, and detection’s dynamic range (12, 13) over microarrays, we characterize the gene expression signature of human diseases through differential expression and functional enrichment analyses (*SI Appendix,* Fig. S1). Then, we construct a Disease Similarity Network (DSN) based on the correlations between diseases’ differential expression profiles. Afterwards, we build a Stratified Similarity Network (SSN) grouping patients of the same disease with a similar expression profile (here called meta-patients). Both networks are able to significantly provide a sizeable coverage (up to 64%) of the medically known disease co-occurrences (2), furnishing a well-defined set of genes and molecular functions potentially implicated in comorbidities, which can be analyzed at http://disease-perception.bsc.es/rgenexcom/ (Fig. 1).

**Figure 1.**
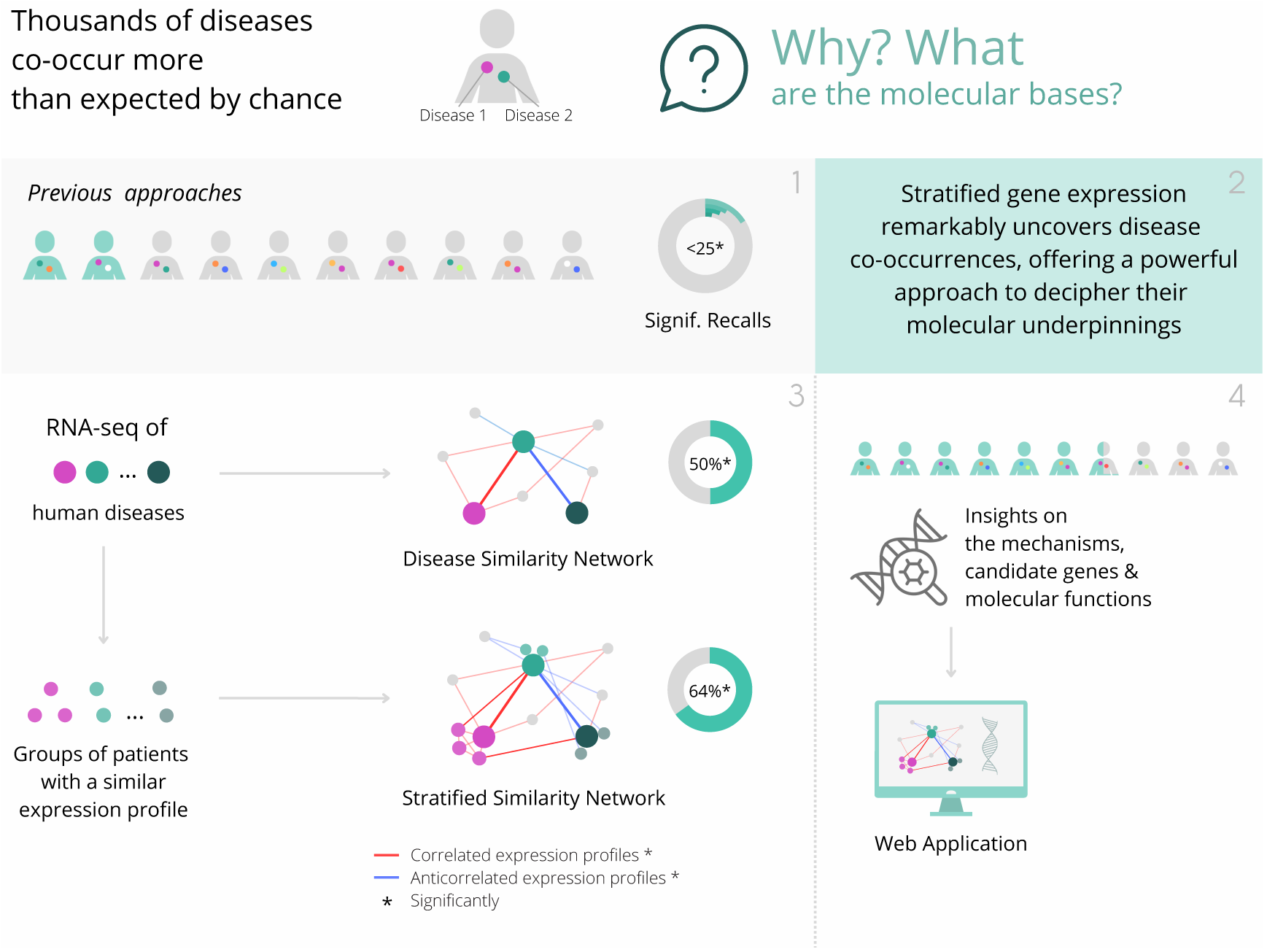
Graphical summary of the study. We build disease and stratified similarity networks that significantly uncover a substantial proportion of disease co-occurrences, providing a powerful approach to decipher their molecular underpinnings.

## Materials and Methods

### Gene Expression Analysis

Uniformly processed RNA-seq gene counts were downloaded from the GREIN platform (14) for 72 human diseases analyzed by 107 studies, including a total number of 4.267 samples. An RNA-seq pipeline destined to the parallel processing of a collection of RNA-seq studies for a given set of diseases was developed (*SI Appendix,* Fig. S2). First, samples with a percentage of aligned reads to the genome lower than 70% were removed, as well as studies with less than 3 cases (from now on patients) and control samples meeting the mentioned requirement. Secondly, gene counts and metadata for each disease were integrated (only studies with the disease, tissue and disease state information were considered). We performed quality controls using the edgeR pipeline (15) and we applied within-sample normalization by considering the logarithm of the counts-per-million (log2CPM). Afterwards, we filtered out lowly expressed genes (those with less than 1 log2CPM in more than 20% of the samples) and we applied between-sample normalization using the trimmed mean of M values (TMM) method (16). After performing batch effect identification, we used the limma pipeline (17) for differential expression analysis. Specifically, we built a model considering sample type (case vs. control) as our outcome of interest and adjusting for the study effect, as it is the most descriptive independent variable (tissue, platform and others depend on the study of origin). Genes with an FDR≤0.05 were considered significantly differentially expressed genes (sDEGs). Moreover, we used Combat (18) and QR Decomposition (17) batch effect removal methods to check the clustering of the samples with t-Distributed Stochastic Neighbor embedding (tSNEs) (19).

### Functional enrichment analysis

In order to better characterize the molecular processes underlying the analyzed diseases, we performed Gene Set Enrichment Analyses (GSEA) (20) on the ranked lists of genes derived from differential expression–log Fold Change (logFC)–using annotations from Reactome (21), Kyoto Encyclopedia of Genes and Genomes (22) and Gene Ontology (23). Gene sets and pathways with an FDR≤0.05 were considered significant.

To enhance the interpretation of altered molecular processes in diseases and their potential involvement in comorbidity relationships, we selected the Reactome pathways significantly enriched in each disease. Subsequently, we applied Ward2 algorithm (24) to cluster diseases based on the Euclidean distance of their binarized Normalized Effect Sizes (1s and-1s representing up-and down-regulated pathways, respectively) (Fig. 2). This clustering approach aids in revealing patterns of shared molecular alterations among diseases, contributing to a more nuanced understanding of potential comorbidity relationships.

**Figure 2.**
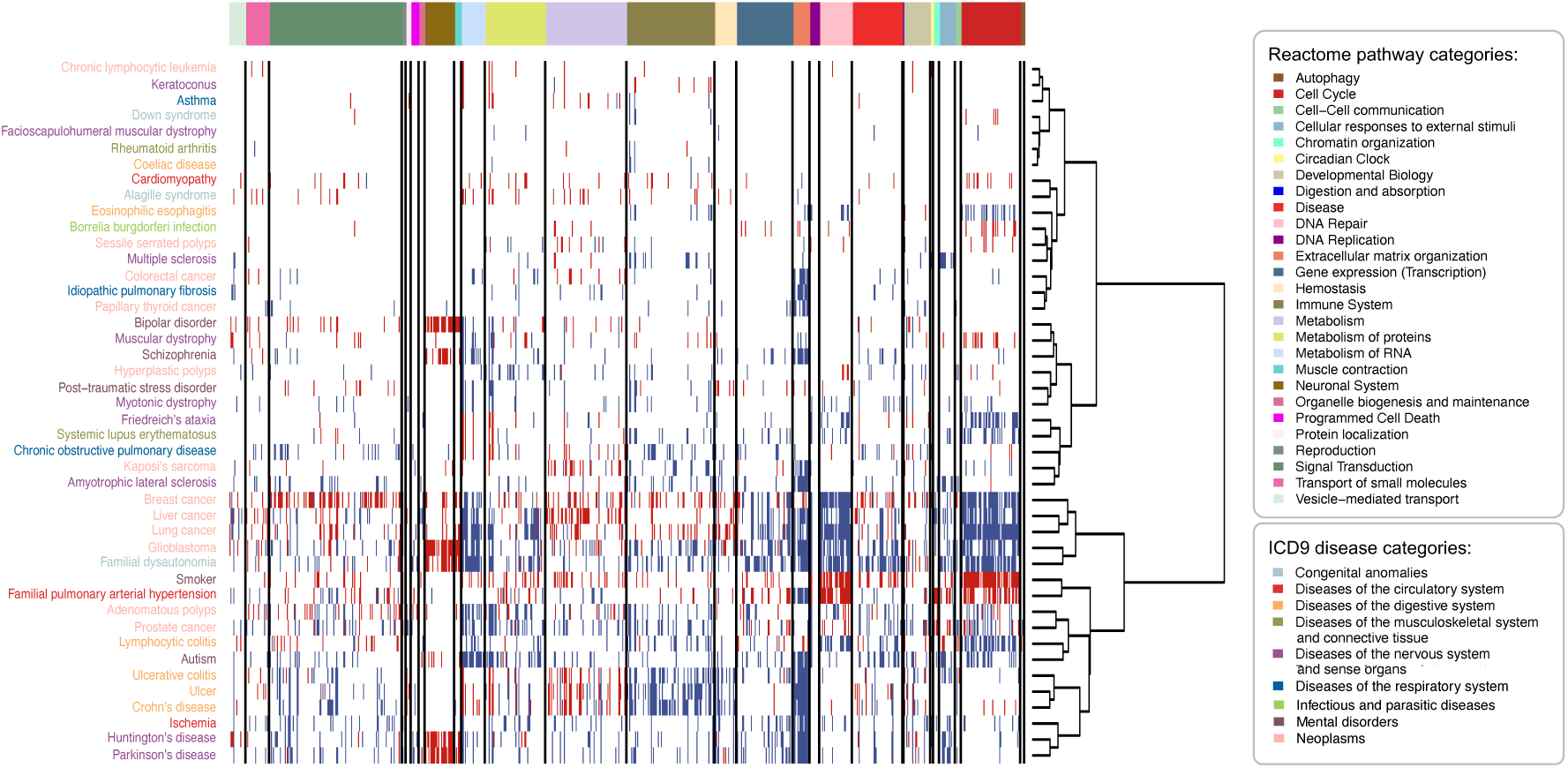
**Reactome pathways significantly altered in human diseases, grouped by pathway category**. For each disease, Reactome pathways significantly over and underexpressed were identified using the GSEA method (FDR≤0.05). Ward2 algorithm was applied to cluster diseases based on the Euclidean distance of their binarized Normalized Effect Size (1s and-1s for over and underexpressed pathways). The heatmap shows the dysregulated Reactome pathways (rows) in the diseases (columns), with over and underexpressed pathways denoted by blue and red colors. Reactome pathways are sorted and grouped by pathway categories, separated by black horizontal lines. Diseases are color-coded based on ICD9 disease categories. Diseases with no dysregulated pathways are not displayed.

### Disease similarity network

To define disease-disease similarities we computed, for each disease pair, the Spearman’s correlation between the logFC values of the genes in the union of their sDEGs. We kept the interactions between different diseases that were significant after multiple testing correction (FDR≤0.05). The resulting Disease Similarity Network (DSN) has the Spearman’s correlation as the edge’s weight, encompassing both positive and negative interactions. Consequently, we computed the topological properties separately for the unweighted positive and negative subnetworks.

Then, we evaluated the overlap of the positive interactions extracted from diseases’ gene expression similarities with those described by Hidalgo et al. (2), based on medical records. To do so, we transformed our disease names into the International Classification of Diseases, Ninth Revision (ICD9 codes). Since some diseases share the same three-digits ICD9 code (e.g. muscular dystrophy, myotonic dystrophy and facioscapulohumeral dystrophy share the code 359-muscular dystrophies and other myopathies), we grouped their samples together and ran the entire analysis (gene expression analysis and disease-disease network building) on them, generating an ICD9 similarity network. Next, we computed the overlap as the percentage of interactions from the epidemiological network–entailing common diseases–captured by ICD9 DSN’s positive and negative interactions independently (recall). To show the network’s enrichment in epidemiological interactions, we computed the overlap in the opposite direction. That is, the percentage of ICD9 DSN interactions contained in the epidemiological network (precision). We assessed the significance of the overlaps by shuffling the interactions while preserving the degree distribution (*SI Appendix,* Note S1). This evaluates if the obtained recalls and precisions significantly surpassed those expected by chance in networks of analogous size and structure. We also computed the overlaps directly from the DSN (*SI Appendix,* Note S2, Table S1). We computed these overlaps with the epidemiological network based on relative risks (RR) and compared them with those obtained using the epidemiological network from Hidalgo et al. based on phi-correlation–a complementary comorbidity measure particularly suited to uncovering true interactions between diseases with similar prevalence that may be missed by RR (*SI Appendix,* Note S3). We performed saturation analysis to evaluate the robustness of the results to different minimum sample size thresholds. Specifically, we systematically assessed how increasing sample size thresholds affect (1) the association between sample size and number of sDEGs or node degree, and (2) the overlap with the epidemiological network based on phi-correlation.

We compared the topological properties of the positive ICD9 DSN and the epidemiological network over the common set of diseases. We computed the number of connected components (to assess network cohesion), mean degree (average connections per node, indicating network density), mean distance (average shortest path length between nodes, indicating network efficiency), density (ratio of actual to possible connections, reflecting network compactness), and diameter (longest shortest path, showing network reach). Further, we calculated mean transitivity (the probability that two nodes sharing a common neighbor are connected, illustrating network cliquishness), mean closeness (how rapidly information traverses from a node to the rest of the network), mean betweenness (mean frequency of a node occurring on the shortest paths between other nodes, indicating centrality), and mean degeneracy (network resilience to node removal). Finally, we evaluated the tendency for diseases to connect with others in the same disease category by computing the assortativity of the network labeling the nodes according to their ICD9 classification. The edge weight was not considered for these calculations. Paired t-tests were employed, where appropriate, to determine the statistical significance of differences in these topological metrics between the two networks, under the null hypothesis that their means are equal.

Next, we compared our overlap with that obtained from other disease-disease networks based on molecular data. We downloaded networks linking diseases based on the similarities of their microbiome (10), miRNAs (9), subcellular localization (11) and cellular components (25). Additionally, we generated a disease-disease network based on protein-protein interactions (PPIs) by selecting the disease pairs that present a significant overlap of their network modules as described by Menche et al. (6). Next, we computed the overlap of these networks with the epidemiological network from Hidalgo et al. (2) over their shared set of diseases and the subset common to our disease set. Finally, we compared the ICD9 DSN with the networks that showed the largest significant overlap with the epidemiology; that is, the network based on PPIs (6) and the microarrays’ disease molecular similarity network by Sánchez-Valle et al. (8).

### Meta-patients generation

We stratified diseases into subgroups of patients with similar expression profiles (meta-patients) by applying clustering algorithms to the normalized and batch effect corrected gene expression matrix. Both PAM (k-medoids) (26) and Ward2 (24) algorithms were applied independently.

In the k-medoids approach, we calculated pairwise distances as 1 - the Spearman’s correlation. To obtain the diseases’ meta-patients, we first determined the optimal number of clusters for each disease by running k-medoids for cluster numbers ranging from 2 to 15. After that, we selected the cluster number with the highest average Silhouette value to obtain the final meta-patients (27).

To evaluate our approach, we selected breast cancer, a disease with known molecular subtypes and for which we have two independent studies. We compared our two independently obtained clusters with the predefined disease subtypes.

### Stratified Similarity Network

To analyze the disease subtype-associated comorbidities, we built the Stratified Similarity Network (SSN) connecting meta-patients and diseases based on the pairwise Spearman’s correlation of the union of their sDEGs. First, meta-patient’s gene expression analysis was performed using the same approach described for the diseases, where all the samples corresponding to a given meta-patient were compared with all controls for the disease. Then, the SSN was built following the same methodology described for the DSN by treating meta-patients and diseases equally as phenotypes. Thus, the SSN contains two types of nodes (diseases and meta-patients) and all the significant interactions between them; i.e. interactions between diseases, between meta-patients, and connections between diseases and meta-patients.

To evaluate if the inclusion of meta-patients significantly enhances detection power, we generated 1000 random meta-patients for each disease by shuffling cases while maintaining the number and size of meta-patients. Next, we obtained 1000 SSNs and assessed whether the number of positive and negative interactions in the SSN could be observed by chance. To evaluate if meta-patients capture epidemiological associations with diseases, we selected the positive interactions between meta-patients and diseases, transformed them into ICD9 codes and computed their overlap with the epidemiological network from Hidalgo et al., as described for the DSN. This is comparable to the available epidemiological network, that comprises interactions at the disease level by evaluating if a group of patients from a given disease is at a higher risk of developing a specific secondary condition. Since epidemiological networks can lack recently validated disease co-occurrences, we performed a literature-based validation for a considerable subset of the obtained interactions (*SI Appendix,* Note S4).

### Web application

To enhance the visualization and exploration of the generated networks, we developed a web application that dynamically displays both the DSN and SSN (28). This interactive tool allows users to filter the networks based on the type of interactions (positive or negative) and set minimum and maximum thresholds for edge weights. Users can also derive network backbones using metric and ultra-metric closures from Simas et al. (29) (*SI Appendix,* Note S5, Table S3). Community detection algorithms such as greedy modularity optimization (30) or random walks (31) can be applied to the filtered networks and interactions involving specific nodes, genes and pathways from Reactome, KEGG, and GO can be selectively filtered. Furthermore, the application facilitates easy inspection and comparison of molecular mechanisms behind diseases and their interactions, enabling users to gain valuable insights into the complexity behind and between diseases at a molecular level.

## Results

### Gene expression fingerprint of human diseases

We first collected publicly available RNA-seq studies focused on human diseases. Uniformly processed gene counts were obtained from the GREIN platform (14). After quality filtering, 58% of the samples were kept, corresponding to 2.705 samples from 62 studies and comprising 45 diseases (Methods, Dataset S1).

We performed differential expression analyses to obtain significantly differentially expressed genes (sDEGs) for each disease (Methods, *SI Appendix,* Table S4). As expected, the number of sDEGs positively correlates with the sample size, whereas it is not affected by the sequencing depth of the diseases, measured by the average library size (*SI Appendix,* Fig. S3).

To functionally characterize the transcriptomic alterations across diseases, we performed functional enrichment analyses (20). We selected the significantly enriched Reactome pathways (FDR≤0.05) (21) and clustered diseases based on their effect sizes (Methods, Fig. 2). For instance, diseases of the digestive system like the Inflammatory Bowel Diseases (IBD): Crohn’s disease and ulcerative colitis, as well as the closely related lymphocytic colitis and ulcer, present an overexpression of a broad set of immune system-related pathways (*SI Appendix,* Fig. S4a). Particularly, Crohn’s disease, ulcerative colitis, and ulcer form a distinct cluster, sharing pathways primarily related to cell-cell communication, hemostasis, metabolism, and signal transduction (*SI Appendix,* Fig. S4b-d, S5a). Additionally, a common overexpression of multiple extracellular matrix (ECM) organization pathways is observed in IBD (*SI Appendix,* Fig. S6c), recently described not only as a consequence of the *in situ* inflammation but also as an active mediator of it (32). Among others, ECM processes are also observed and described to be altered in diseases for which IBD is a risk factor, such as ulcer and colorectal cancer (33, 34).

### Disease similarity network

Next, we built a Disease Similarity Network (DSN) connecting diseases based on the significant correlation of their differential gene expression profiles. Specifically, for each disease pair, we considered the Spearman’s correlation (FDR≤0.05) of the fold changes of the genes within the union of their sDEGs (Methods, Fig. 3). The resulting DSN forms a single connected component, comprising 63.37% positive interactions and 36.63% negative interactions, with a mean degree of 29.24 (*SI Appendix,* Table S5). Positive interactions indicate a positive correlation between the differential gene expression profiles of the diseases, while negative interactions denote an inverse correlation (*SI Appendix,* Note S6). Additionally, node degree positively correlates with both the number of sDEGs and the sample size of the diseases, suggesting that diseases with more sDEGs and larger sample sizes tend to have a higher number of connections in the network (*SI Appendix,* Fig. S7, Note S7).

**Figure 3.**
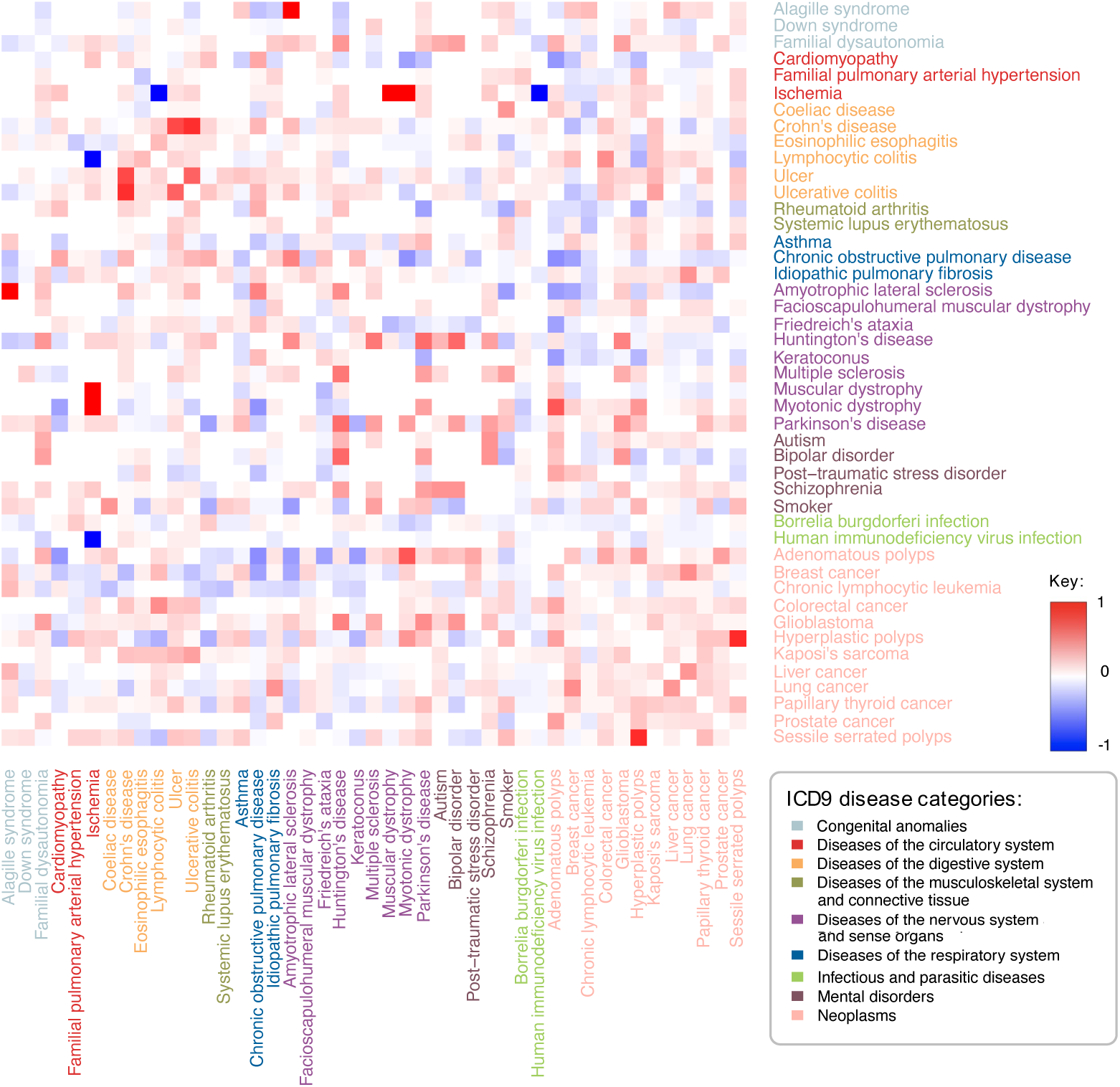
Heatmap representation of the Disease Similarity Network. Pairwise disease correlations were computed based on the Spearman’s correlation of the union of the sDEGs of each pair of diseases. Subsequently, a disease-disease network was built, containing the significantly positive and negative correlations (FDR≤0.05), with edge weights corresponding to Spearman’s correlations (Methods). The heatmap shows the positive and negative disease interactions in red and blue, respectively. White indicates pairs of diseases that are not connected. Diseases are color-coded based on ICD9 disease categories.

A key strength of this network is its capacity to reflect both direct and inverse comorbidities– diseases that co-occur significantly more or less than expected by chance, respectively. For example, the DSN captured the previously described IBD comorbidities, as well as intra-disease category comorbidities regarding neoplasms (e.g. lung and liver cancer) (Fig. 3). We also observed positive correlations between seemingly disparate conditions, such as Kaposi’s sarcoma and human immunodeficiency virus’ infection (HIV), being this neoplasm one of the most common malignancies in HIV patients as a consequence of their immune deficiency (35). Furthermore, the network unveiled less frequent associations, such as those observed between Kaposi’s sarcoma and other diseases characterized by immune system imbalances, like IBD (36, 37).

Many positive interactions entailing diseases of the nervous system and mental disorders are observed, mainly due to shared neurological dysfunction, ECM dysregulation and, in some cases, immune system involvement (Fig. 2-3). Among others, schizophrenia was found to be connected to bipolar disorder, autism, and Huntington’s and Parkinson’s diseases, which are known to be comorbid.

Conversely, we observed some negative correlations between the expression profiles of specific central nervous system disorders (CNSd) and cancers (38), reflecting divergent molecular profiles. For instance, Huntington’s disease exhibits a negative interaction with liver, lung, and breast cancer and chronic lymphocytic leukemia, which are known to co-occur less than the expected by chance (39). Upon examining the altered pathways in these diseases, we discovered opposing molecular tendencies in many of their key pathogenic processes, with approximately 85% of shared altered pathways showing opposite regulation. On one hand, cancer is characterized by an overexpression of cell cycle and gene transcription mechanisms, whereas Huntington’s disease shows increased cell death, apoptosis, mitochondrial dysfunction, and a negative regulation of gene transcription (*SI Appendix,* Fig. S5c-d, S6a). Pathways related to kinesins, involved in cell division and intracellular transport, are overexpressed in cancer as previously described (40), while underexpressed in Huntington’s disease, where its impairment is also characterized (41). In addition, immune abnormalities have been extensively documented as central to both Huntington’s disease and cancerous processes (39, 42). For example, we observed increased interleukin production and signaling regarding Th1-type immune response (e.g. IL-12) in Huntington’s disease, alongside activation of the complement cascade. Indeed, these processes are underexpressed in cancer and have been previously linked to carcinogenesis (*SI Appendix,* Fig. S4a, Note S8) (42–44).

Subsequently, we evaluated to what extent the DSN is able to capture medically known comorbidities by computing its overlap with the epidemiological network from Hidalgo et al., based on relative risks (RR) (2) (*SI Appendix,* Note S3). We observed that the DSN significantly recalls 46.2% of the interactions in Hidalgo et al. over the common set of diseases (p-value=0.0018), underscoring the DSN’s capability to mirror known disease relationships (Methods, *SI Appendix,* Table S6). Since epidemiological networks lack reliable estimations of inverse comorbidities, negative interactions in the DSN could not be systematically evaluated. Despite this, our analysis aligned with expectations, showing a non-significant overlap of these negative interactions with direct comorbidities (p-value=0.867). To ensure robustness, we built a DSN using an alternative non-linear metric (cosine similarity) and showed that very similar and even better results were obtained (49.25% recall, p-value=0.0034) (*SI Appendix,* Table S7). Moreover, the cosine-based DSN is enriched in the interactions obtained with Spearman, for both positive (Odds ratio=3.3, p-value=2.2e-16) and negative (Odds ratio=5.52, p-value=2.2e-16) links (Fisher’s exact test). Robust recall and markedly improved precision are obtained by considering an alternative epidemiological network from Hidalgo et al. based on phi-correlation, which accurately detects comorbidities between diseases with similar prevalence that may be missed by the RR (*SI Appendix,* Table S8). These results indicate that a substantial proportion of the initially predicted false positives actually reflect true disease co-occurrences. Additionally, we confirmed that meaningful and biologically relevant disease co-occurrences are effectively captured even for diseases with limited sample sizes, such as rare diseases, with some of them even exhibiting above average performance (e.g. Kaposi’s sarcoma or familial dysautonomia) (*SI Appendix,* Note S9).

The DSN’s precision and recall varies depending on the disease category pair (Fig. 4). Diseases of the digestive system present the highest precision (66.4%) with a mean recall of 53.6%. Interactions entailing congenital anomalies are also captured at a high level. On the contrary, highly heterogeneous diseases (e.g. mental disorders) tend to present lower recall values, and neoplasms, which often share the dysregulation of multiple pathways without being comorbid, exhibit the lowest precision.

**Figure 4.**
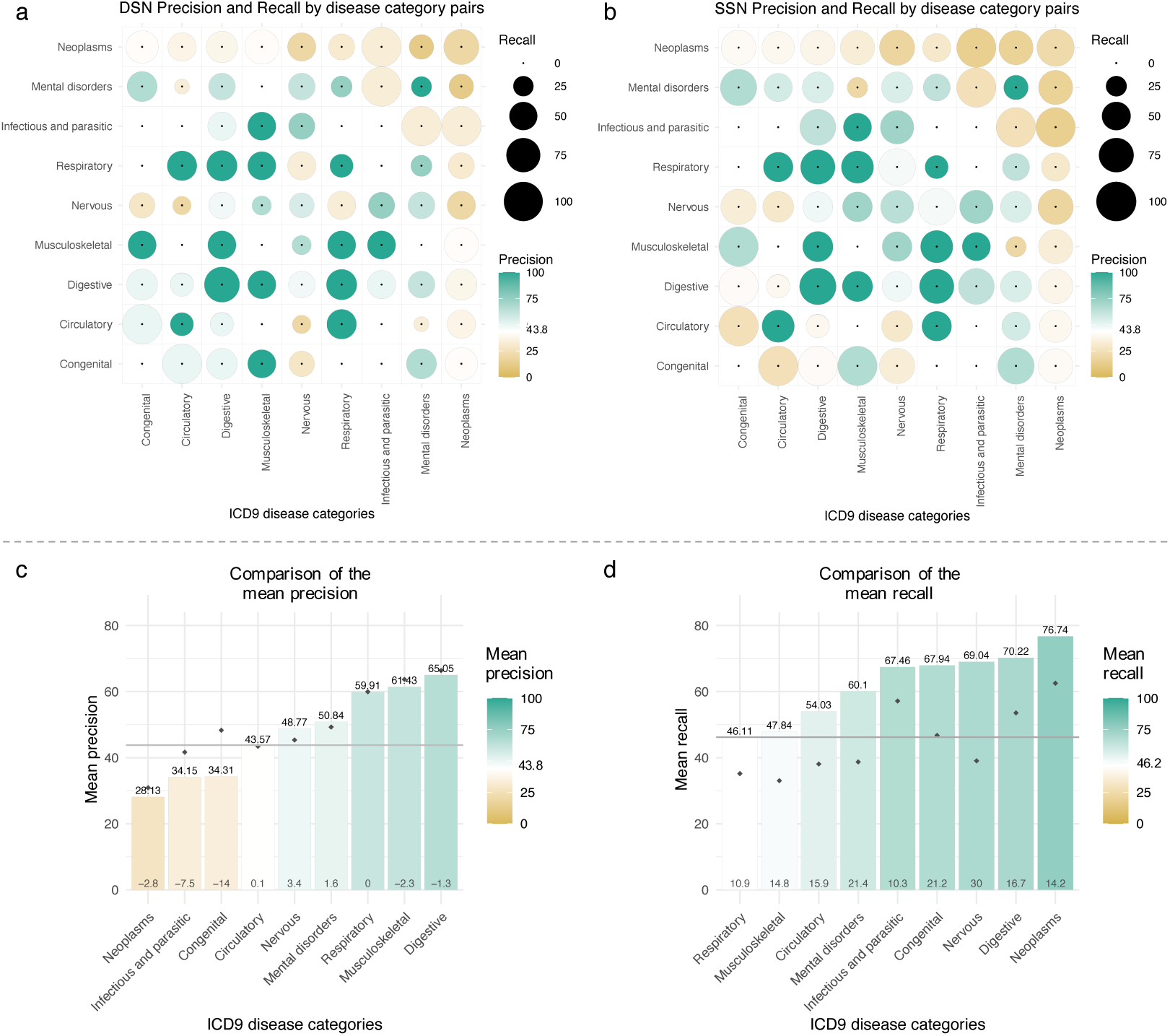
Precision and recall of the DSN and SSN by disease category. Precision and recall of the **(a)** Disease Similarity Network (DSN) and the **(b)** Stratified Similarity Network (SSN) by disease category pairs. The precision is the percentage of interactions in the molecular networks present in the epidemiological network from Hidalgo et al. for the common set of diseases. The recall is the percentage of epidemiological interactions captured by the molecular networks for the common set of diseases. For the SSN, interactions between meta-patients and diseases were considered (Methods). Each point in the symmetric matrix corresponds to the subnetwork resulting from selecting interactions between diseases of the indicated disease categories. Circle area represents recall, and color corresponds to precision. Green and yellow colors indicate higher and lower precisions than the one of the DSN (43.8%), shown in white. Disease pairs without epidemiological interactions present a single black point. **(c)** Mean precision of each disease category in the SSN. Disease categories are sorted by mean precision. Green and yellow indicate higher and lower precisions than the one of the DSN, represented by the horizontal grey line. **(d)** Mean recall of each disease category in the SSN. Disease categories are sorted by mean recall. Green and yellow indicate higher and lower recalls than the one of the DSN (46.2%), represented by the horizontal grey line. Black points indicate the mean **(c)** precision or **(d)** recall of the disease categories in the DSN. Differences between meta-patient and disease-level values are plotted at the base of the bars.

In addition, we found that the DSN and the epidemiological network share remarkably similar structural makeup, despite their differing origins and nature of their underlying data. Both networks comprise a single connected component and exhibit closely matched density and mean distance, with values around 0.42 and 1.6, respectively, underscoring comparable connectivity and navigational efficiency across the networks (Methods, *SI Appendix,* Table S9). Furthermore, the networks show no significant differences in key topological metrics such as mean degree, closeness, betweenness, and degeneracy, indicating a resemblance in their overall structure, density, centrality, node removal resilience, and information flow. Both networks display a neutral preference for within-disease category links, which are slightly more frequent in the DSN (assortativity, Methods) (*SI Appendix,* Note S10). Even with some small differences, the observed substantial structural congruence between the networks supports the robustness of the DSN in mirroring complex disease relationships.

Next, we compared our overlap with those derived from published disease-disease networks based on other molecular data (Methods). Interestingly, these approaches disclosed significant correlations or enrichments with epidemiological networks but did not quantify their recall with them, except from (8). Networks based on the microbiome (10), miRNA (9) and subcellular localization similarities (11) yielded non-significant overlaps with the epidemiology over their respective common diseases and over the common diseases also present in the DSN (Table 1). The network derived from protein-protein interactions (PPIs) (6) showed significant yet small overlap with the epidemiological network (8.71% for the entire network and 18.52% over the diseases in the DSN). Similarly, the network generated by Sánchez-Valle et al. (8) using microarrays demonstrated a significant overlap of 16% with the epidemiological network from Hidalgo et al. (2). Likewise, an approach based on cellular components showed significant and very small recalls of 3.3% at the level of shared genes and 6.6% at the level of PPIs between disease genes (25). When considering the diseases in the DSN, only the network built using shared genes presented a significant overlap of 11.22% (Fig. 5a, *SI Appendix,* Table S10).

**Figure 5.**
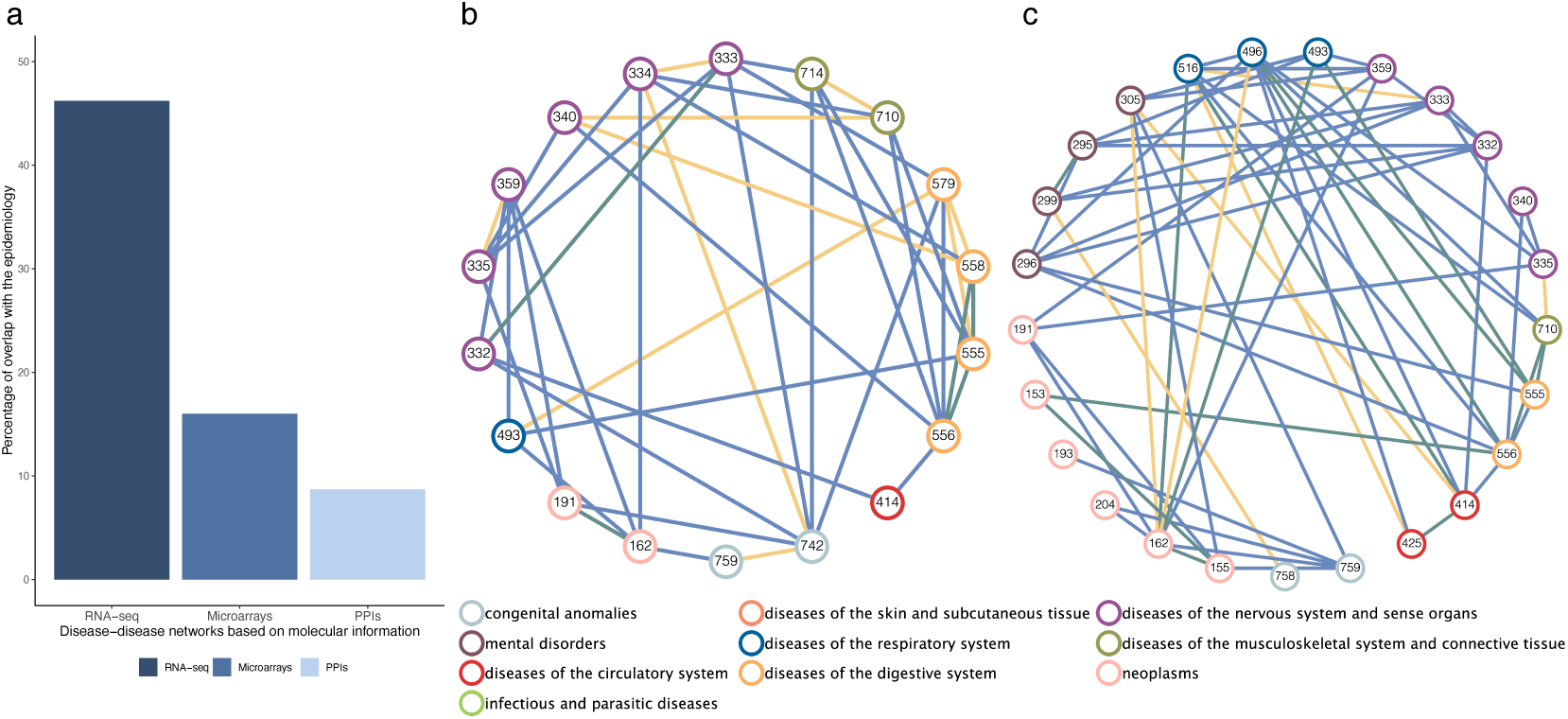
Comparison of the epidemiological interactions described in the Disease Similarity Network (DSN) and other molecular networks. **(a)** Percentages of interactions in the epidemiological networks, significantly captured by molecular networks (recall). It shows the recall of the top three approaches; that is, our DSN based on RNA-seq, the network based on microarrays, and the approach based on protein-protein interactions (PPI) (Methods). **(b)** Network visualization of the positive disease-disease interactions described in the epidemiological network by Hidalgo et al. captured only by the DSN (blue), only by the network based on PPIs (6) (yellow), or by both (green). It shows the interactions over the common set of ICD9 codes in both molecular networks, with diseases colored by their ICD9 category. **(c)** Network visualization of the positive disease-disease interactions described in the epidemiological network by Hidalgo et al. captured only by the DSN (blue), only by the network based on microarrays (8) (yellow) or by both (green).

**Table 1.** Recall of the disease-disease networks based on molecular information with the epidemiology. Table showing the recall and p-value of the Disease Similarity Network (DSN), the Stratified Similarity Network (SSN), and other disease-disease networks based on molecular information with the epidemiological network from Hidalgo et al. (2). The assessment is conducted over the entire networks at the ICD9 level (All ICD9) and the ICD9 codes considered in the presented analysis (ICD9s in the DSN). Asterisks indicate that the recall is significant.

|  |  | All ICD9 |  | ICD9 in the DSN |  |
| --- | --- | --- | --- | --- | --- |
|  | Network | Recall (%) | p-value | Recall (%) | p-value |
| Urda-García et al. | DSN | 46.2* | 0.0018 | 46.2* | 0.0018 |
|  | SSN | 64.13* | 0.0187 | 64.13* | 0.0187 |
| Previous networks | Microbiome | 36.09 | 0.067 | 63.64 | 1 |
|  | miRNA | 36.03 | 0.412 | 56.25 | 0.96 |
|  | PPIs | 8.71* | 0 | 18.52* | 0.0045 |
|  | Cellular components (PPIs) | 6.63* | 9e-4 | 26.53 | 0.2 |
|  | Cellular components (genes) | 3.26* | 0 | 11.22* | 0.0095 |
|  | Subcellular localization | 23.64 | 0.269 | 36.45 | 0.639 |

Finally, we compared the molecular networks with the highest significant overlap (PPI and microarray-based networks) to the DSN (*SI Appendix,* Table S11-12). As figure 5b-c shows, most comorbidities involving common diseases are only captured by the DSN, which matches over 140 comorbidities not captured by each of these approaches. Interestingly, the DSN significantly recalls 62.22% (p-value=0.002) of the positive interactions in the microarray’s network (*SI Appendix,* Note S11).

### Molecular mechanisms behind comorbidities

Once confirmed that the DSN captures a significant percentage of comorbidities, we delved into the molecular mechanisms behind its epidemiological (EIs) and non-epidemiological interactions (NEIs), i.e. interactions without a clear correspondence with current medical data. We identified the Reactome pathways commonly over and underexpressed for each pair of diseases, grouping them by Reactome pathway category. We observe that the pathway categories associated with most interactions tend to display a broader range of dysregulation, involving a higher number of altered pathways (Fig. 6). Impressively, 95.2% of the EIs in the DSN share at least one–and a mean of 21.2–overexpressed immune system pathways, underscoring the potentially pervasive role of the immune system in disease co-occurrences (*SI Appendix,* Note S12, Table S13). Additionally, pathways related to the ECM, metabolism of proteins, metabolism and signal transduction are involved in over 90% of the interactions, with means of 10.9, 10.1, 6.3, and 16.1, respectively. On the other hand, underexpressed pathways associated with most EIs are primarily linked to the metabolism of proteins, signal transduction, metabolism, and developmental biology with means of 4.9, 7.2, 13.5, and 2.5, respectively.

**Figure 6.**
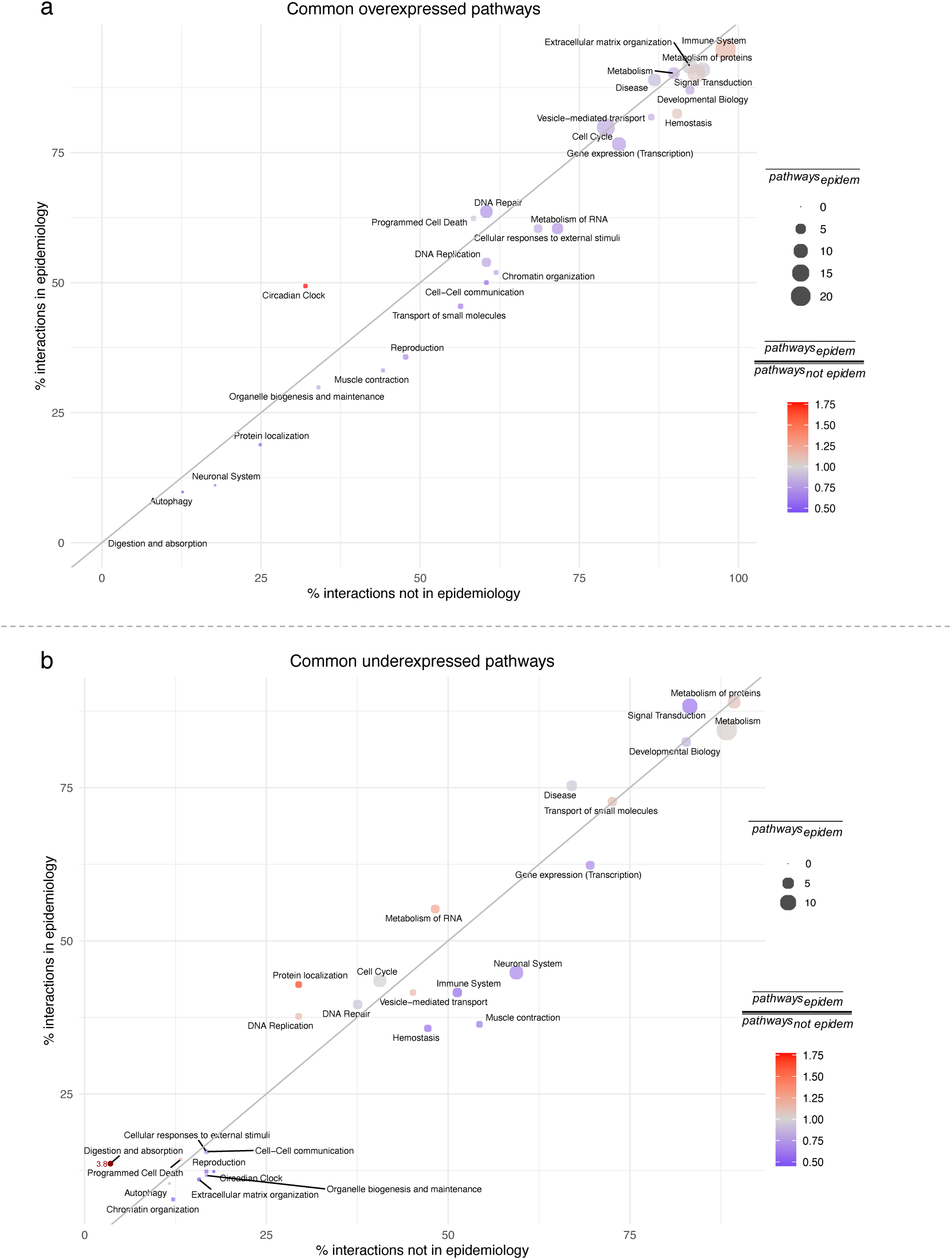
Over and underexpressed pathways behind epidemiological and non-epidemiological interactions. Percentage of epidemiological versus non-epidemiological interactions sharing **(a)** overexpressed or **(b)** underexpressed pathways. Each data point represents a Reactome pathway category, with point size indicating the mean number of shared pathways in epidemiological interactions. Point color reflects the ratio of the mean number of shared pathways in epidemiological versus non-epidemiological interactions (e.g. red indicates that epidemiological interactions share more altered pathways than non-epidemiological interactions).

To adequately detect which pathways are specific to EIs, we also considered the ratio between the mean number of shared over or underexpressed pathways by Reactome category in EIs versus NEIs (Fig. 6, *SI Appendix,* Fig. S8). Overexpressed circadian clock pathways explain more EIs than NEIs, with the mean number of shared pathways being 1.7 times higher in the former. Digestion and absorption and protein localization are 3.8 and 1.6 times more commonly underexpressed in EIs, suggesting their repression or inactivation across co-occurring diseases.

Higher resolution results can be obtained by considering interactions between pairs of disease categories. For instance, the shared overexpression of circadian clock pathways seems to be highly specific of EIs entailing CNSd (Fig. 6, 7a). Actually, the pivotal and putatively causal role of the circadian system in CNSd and their comorbidities has recently been proposed (45–47). Although each individual comparison has its particular portrait of the mechanisms underlying disease interactions, some general patterns become apparent. We observe that pathways tend to cluster according to their ability to explain EIs versus NEIs. In Fig. 7a we can see a cluster of pathways whose dysregulation is shared by all EIs, whereas smaller clusters are mostly present in NEIs. Interestingly, pathway categories involved in more EIs than NEIs also present a higher number of pathways commonly dysregulated in EIs than in NEIs (Fig. 7a, within the upper cluster, redder dots are observed at the left side). In summary, EIs have been found to present more shared altered pathways than NEIs. In fact, if we remove neoplasms, EIs share the alteration of 53.1% and 56.8% more over and underexpressed pathways, respectively. This highlights a closer molecular relationship between co-occurring diseases, supporting the evidence of molecular mechanisms driving their co-occurrence.

**Figure 7.**
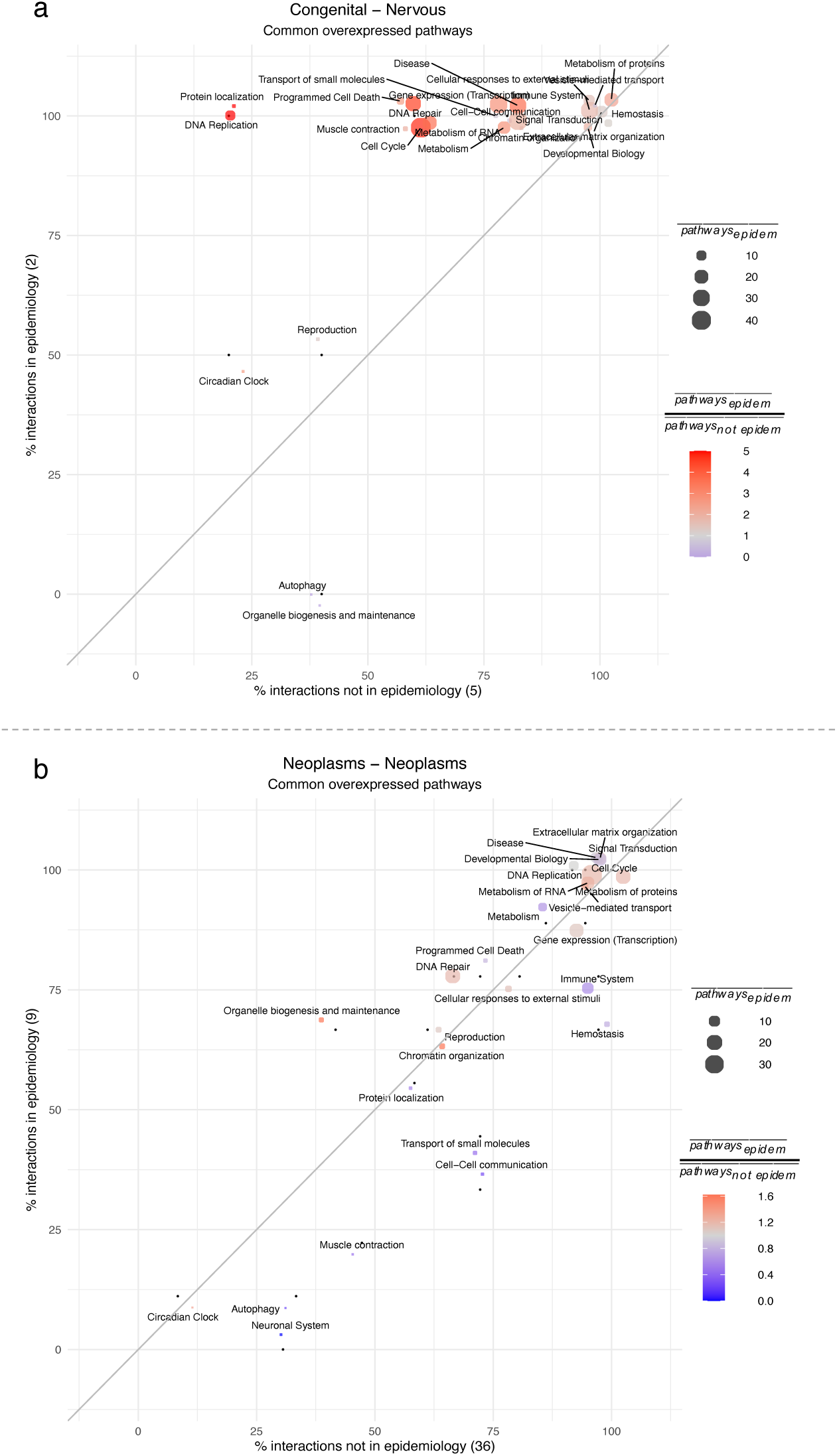
Examples of overexpressed pathways behind epidemiological and non-epidemiological interactions. Percentage of epidemiological (EIs) versus non-epidemiological interactions (NEIs) that share overexpressed pathways. Each data point represents a Reactome pathway category, with point size indicating the mean number of shared overexpressed pathways in EIs. Point color reflects the ratio of the mean number of shared pathways in EIs versus NEIs (e.g. red indicates that epidemiological interactions share more pathways than non-epidemiological interactions). The number of EIs and NEIs is indicated in parentheses on the y and x axis labels, respectively. **(a)** Interactions between congenital anomalies and diseases of the nervous system and sense organs. **(b)** Interactions within neoplasms.

By examining the interactions between neoplasms, we can discern between the mechanisms that are potentially responsible for their common cause from those more likely due to the convergence of some common functions (i.e. overexpression of pathways related to the immune system or cell-cell communication as well as the underexpression of developmental biology or DNA repair processes) (Fig. 7b, *SI Appendix,* Fig. S9). Comorbid neoplasms tend to share a higher number of overexpressed pathways related to organelle biogenesis and maintenance, chromatin organization or cell cycle, along with underexpressed pathways related to metabolism, transport of small molecules or immune system. Interestingly, around 30% of comorbidities within neoplasms share a highly specific underexpression of protein localization pathways.

### Defining disease meta-patients

It has been shown that patients suffering from a given disease often present different risks of developing specific secondary conditions (4). We propose that these differential risks might be driven by the existence of disease subtypes. To explore this hypothesis, we conducted a stratified analysis of disease similarities by applying clustering algorithms to the processed gene expression counts. This enabled us to identify subgroups of cases with a similar expression profile for each disease, which we refer to as meta-patients (Methods). To evaluate our approach, we selected breast cancer, a disease with well-known molecular subtypes for which we have two independent studies. The first study comprises 20 estrogen positive (ER+) samples and 18 triple negative (TN) and the second study includes 9 estrogen negative (ER-) samples and 9 ER+. Our two independently obtained clusters (using PAM (48) and Ward2 (24) clustering algorithms separately) yielded similar results, with PAM more accurately grouping cases according to their defined molecular subtypes (*SI Appendix,* Table S2, Fig. S10). Breast cancer patient’s clustering and pairwise similarity shows that PAM clustering classifies most of the cases correctly into estrogen negative (ER-), triple negative (TN) and estrogen positive (ER+) (Fig. 8a), even grouping cases with shared molecular subtypes that belong to two independent studies.

**Figure 8.**
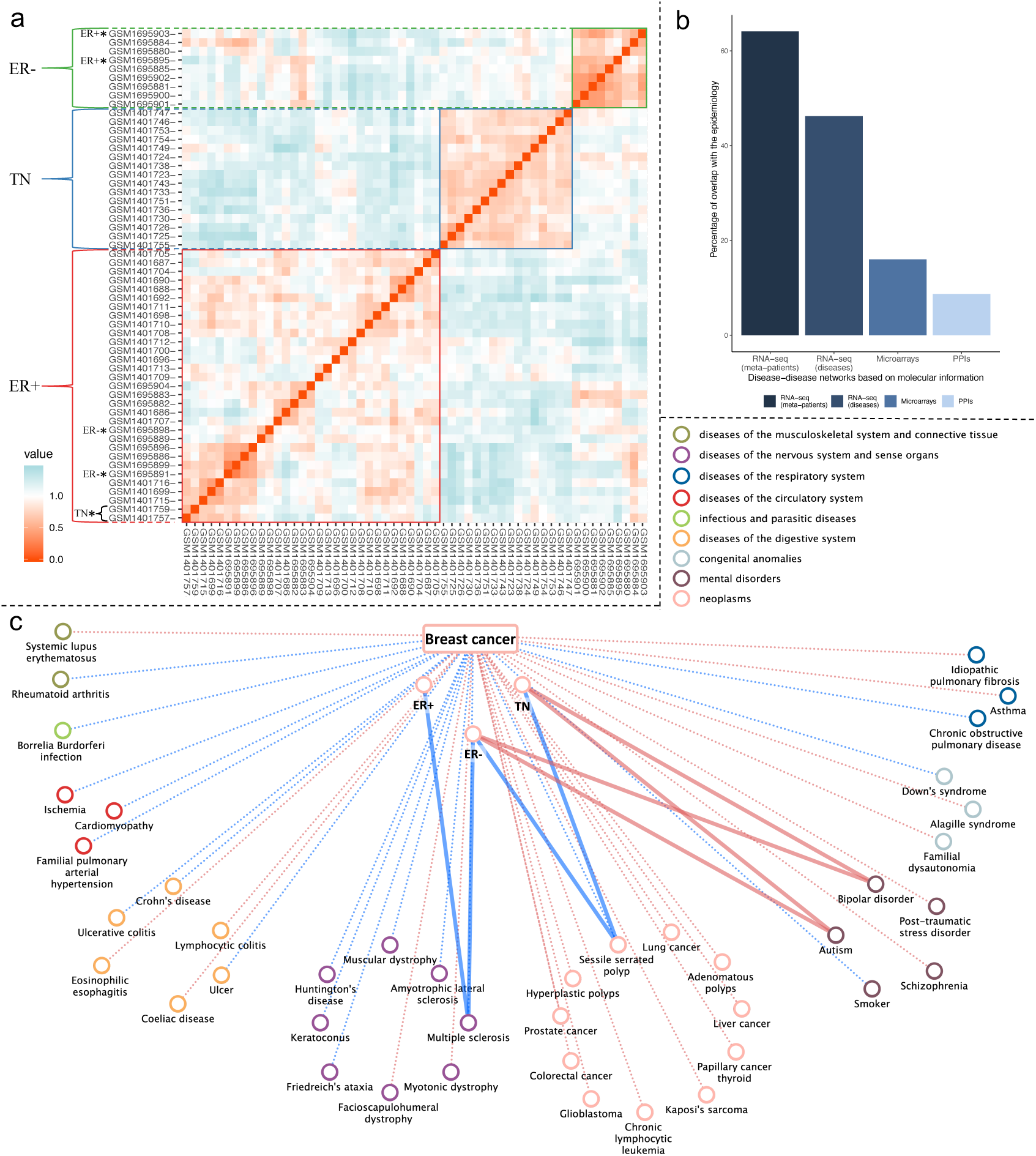
Breast cancer meta-patients and breast cancer molecular similarity interactions at the disease and meta-patient level. **(a)** Heatmap showing breast cancer patients’ similarity and classification through PAM algorithm. Similarity has been defined as 1 – Spearman’s correlation of the gene expression values. Orange denotes a positive correlation, while blue indicates a negative correlation between patient expression profiles. Patients are colored-coded based on their molecular subtype, with ER-(estrogen negative) in green, TNEG (triple negative) in blue, and ER+ (estrogen positive patients) in red. Patient clustering is marked on the left side. **(b)** Percentage of interactions in epidemiological network significantly captured by molecular networks. It shows the overlap of the interactions at the meta-patient and disease level based on RNA-seq and the networks based on microarrays and protein-protein interactions (PPI) (Methods). **(c)** Interactions between breast cancer disease and its meta-patients with human diseases. Positive interactions are represented in red, negative interactions in blue. Dashed-faded lines depict interactions at the disease level shared by all the meta-patients, while solid lines represent meta-patient specific interactions.

### Stratified Similarity Network

Since meta-patients are groups of patients from a given disease, we can derive their significantly altered genes, pathways and molecular similarities. Hence, and first of all, we characterized the obtained meta-patients in terms of their differential expression profiles (Methods). Then, we expanded the DSN by incorporating meta-patients and their differential gene expression profile’s similarities (Methods). The resulting Stratified Similarity Network (SSN) contains two types of nodes–diseases and meta-patients–and all the significant interactions between them; i.e. interactions between diseases, interactions between meta-patients and interactions connecting diseases and meta-patients.

The SSN can be fully explored in the <u>web application</u> and its topological properties can be found in the *SI Appendix,* Table S14. Since the SSN has a considerably higher number of nodes, we observe a large increase in mean degree with respect to the DSN. As captured by the moderate increase in density and mean transitivity, the meta-patients have added some new interactions that were missed by the DSN, being able to uncover new transitive relationships. These properties, along with a concordant decrease in diameter and mean distance, occur across the entire network and subnetworks entailing positive or negative interactions. In fact, by defining meta-patients we significantly increase the number of diseases linked to each disease by 7.64 diseases (p-value=1.596e-10) and 7.29 (p-value=6.159e-15) for the positive and negative interactions respectively (*SI Appendix,* Fig. S11a). We confirmed that this increase in detection power is significant compared to randomly generated meta-patients (p-value=0 for positive and negative interactions) (Methods, *SI Appendix,* Fig. S12).

Human diseases exhibit varying degrees of heterogeneity, reflected in their clinical symptoms, treatment responses, progression, molecular underpinnings, or associated comorbidities. When examining breast cancer, we observe that while most of the interactions are shared across breast cancer meta-patients, there is a significant number of specific interactions (Fig. 8c). Notably, a negative interaction with multiple sclerosis is exclusive to ER+ and ER-meta-patients, hinting at a distinct immunological landscape that could inform targeted research or therapeutic strategies. Conversely, positive interactions with autism and bipolar disorder are uniquely observed in TN and ER-meta-patients, pointing to shared genetic susceptibility and molecular processes (*SI Appendix,* Note S13).

Actually, the proportion of positive and negative links shared by all the meta-patients from a given disease varies greatly (*SI Appendix,* Fig. S11b). For example, CNSd (e.g. schizophrenia, bipolar disorder, multiple sclerosis or autism), known to be highly heterogeneous, show little consistency in their meta-patients’ interactions. On the other hand, neoplasms–that tend to present a consistent and pronounce alteration of multiple biological processes–seem to present a higher correspondence in their meta-patients’ connections.

To assess the ability of meta-patients to uncover new comorbidities, we computed the overlap of the interactions between meta-patients and diseases with the epidemiological data (Methods). Remarkably, meta-patients significantly captured 64.1% (p-value=0.0187) of the interactions in the epidemiological network from Hidalgo et al. This represents a 17.9% increase in recall compared to the DSN, with only a marginal decrease in precision of 0.7%. To further validate these findings, we computed the overlap with the phi-correlation epidemiological network. The results show consistent recall accompanied by a substantial increase in precision, even slightly exceeding that of the disease-level network (*SI Appendix,* Table S8). As expected, negative interactions do not show a significant overlap (p-value=0.8) (*SI Appendix,* Table S15). As depicted in Fig. 4, most disease categories benefit from the stratification of diseases, since they tend to present a considerable increase in recall often accompanied by a slight decrease in precision. Aligned with previous findings, highly heterogeneous diseases (nervous system and mental disorders) present some of the highest increases in recall (up to 30%); strikingly, also gaining precision. This suggests that meta-patients more accurately reflect true comorbidities, enhancing the number of correctly identified disease links while reducing the frequency of false positives. More moderately, this tendency also occurs for circulatory diseases. Respiratory diseases, which often display a wide range of immune system responses, exhibited a 10.9% increase in recall with maintained precision compared to the DSN. Some of the newly discovered comorbidities with this approach include chronic obstructive pulmonary disease’s relationship with smoking or schizophrenia; coeliac disease’s association with asthma, HIV or lupus, among others; and ten additional comorbidities of muscular dystrophy, such as cardiomyopathy, multiple sclerosis or HIV (*SI Appendix,* Data S2).

Since gold standard epidemiological networks may lack recently validated disease co-occurrences, we manually validated the top and bottom 100 false positive interactions between diseases and meta-patients (Methods). By integrating this literature-based validation with the complementary phi-correlation-based epidemiological network, we found that 60% of the false positives identified using the RR-based network are in fact true positives (∼30% supported by phi-correlation and ∼30% through literature validation) (*SI Appendix,* Note S4). Incorporating this correction yields a final precision of 76.65%, indicating that the majority of the predicted disease associations are supported by strong epidemiological evidence. *SI Appendix,* Notes S14 and S15 explore several of these newly identified disease co-occurrences, proposing novel mechanistic explanations for them and suggesting additional disease co-occurrences with molecular evidence.

Finally, we developed a user-friendly web application (http://disease-perception.bsc.es/rgenexcom/) for convenient exploration of the networks and their underlying molecular mechanisms (Methods).

## Discussion

So far, molecular representations of disease interactions have recalled a limited proportion of the medically known disease co-occurrences, proving insufficient to address the long-standing question of the molecular origin of comorbidities. The generated networks based on RNA-seq profiles offer a convincing and comprehensive answer to this matter, being able to significantly capture and meaningfully explain up to 64% of the known comorbidities. Hence, they render a qualitative difference over previous studies, solidly supporting the key role of molecular mechanisms behind disease co-occurrences in a generalized manner.

Our analysis revealed a strong structural congruence between the Disease Similarity Network (DSN) and the epidemiological network, highlighting the DSN’s effectiveness in mirroring complex disease relationships. This similarity across key topological metrics suggests a consistency in how diseases are interrelated, whether viewed through the lens of molecular biology or epidemiological studies. The DSN showed a slight propensity for diseases within the same category to associate more than observed in epidemiology. For instance, while molecular similarities connect some neoplasms that are indeed comorbid (e.g. liver cancer with glioblastoma, lung, or colorectal cancer), they also link specific cancers that are not epidemiologically linked. This shows that the presence of shared molecular mechanisms does not always translate into an increased relative risk that is observed in the currently limited medical data, as previously described by Park et al. (25). However, we can discern between the mechanisms behind well-established comorbidities from the ones that may be due to an overall molecular similarity, which is especially relevant for neoplasms that share similar dysregulated pathways. Indeed, without considering neoplasms, co-occurring diseases tend to share over 50% more altered pathways than pairs of diseases without clear evidence of a medical relation. Furthermore, we found that many of our molecular similarities correspond to recently discovered comorbidities missed by Hidalgo et al., such as breast cancer co-occurrence with colorectal, thyroid, lung cancer or Kaposi’s sarcoma (49), and between-category associations, like the elevated risk of ulcers in thyroid cancer patients due to radioactive iodine treatment (50).

Several ways in which shared molecular mechanisms can underlie direct comorbidities have been proposed. Essentially, molecular mechanisms can be causally or consequentially altered in a given disease; which, in turn, can contribute to the development of a secondary condition. Thus, molecularly based comorbidities can be explained by the following scenarios: (a) both diseases share the same or correlated causal alterations, (b) the molecular mechanisms altered as a consequence of one disease are associated to the second condition or (c) there is a third condition that increases the risk of developing both of them (1). These schemes are not mutually exclusive and can be combined in complex manners. In fact, the study of direct comorbidities in longitudinal studies has shown disease trajectories that can be explained by an underlying aggravation and accumulation of specific molecular processes, especially in a chronic manner (51). This is the case for the discussed progression of IBD into colorectal cancer (52) and for the highly prevalent disease trajectory that has recently been called metabolic syndrome, including obesity, insulin resistance, diabetes, cardiovascular disease or even cancer (53). These observations supported the central role of the underlying molecular mechanisms in the study of individual diseases and their co-occurrences, to the point where efforts have already been destined to redefining diseases by incorporating both their clinical features and molecular profiles (54).

Disease co-occurrences can be better understood if disease subtypes and patient-specific patterns are taken into account (4). Indeed, previous epidemiological studies have identified comorbidities that depend on the disease subtype (55–57). In line with this, we introduced the concept of meta-patients and the stratified exploration of their molecular similarities with diseases. Meta-patients unraveled a significant mean of around 14 new subgroup-specific disease connections per disease, increasing the detection power of disease similarities (*SI Appendix,* Fig. S11a). This subclassification of diseases based on similarities of the patients’ gene expression profiles can be related to epidemiological observations of comorbidities that depend on patients’ characteristics. In the case of breast cancer, we observed that although the three meta-patients share most of the disease interactions, some specific and interesting ones emerge. For instance, TN and ER-meta-patients are the only ones presenting a positive interaction with autism and bipolar disorder (Fig. 8c). While several studies (58, 59) found no significant correlations, recent epidemiological evidence suggests breast cancer as a comorbidity of autism (60). Indeed, a significant overlap between significantly altered genes and pathways of autism and several cancer types has been reported (61). In addition, bipolar disorder has been associated with increased cancer risk, though the risk of breast cancer appears higher but non-significant (62). We also observed a negative interaction between breast cancer, ER+ and ER-meta-patients with multiple sclerosis (Fig. 8c). Again, opposite tendencies are described in the literature for this connection, where the order of appearance of the disease seems to drive the comorbidity pattern. It has been shown that breast cancer patients are 45% less likely to develop multiple sclerosis (63). Conversely, multiple sclerosis patients face a significantly increased risk of breast cancer, presumably driven by immunosuppression from associated treatments (64). Therefore, our analysis provides new evidence on subgroup-specific comorbidities, offering molecular explanations that could inform the development of targeted therapies (*SI Appendix,* Note S13-15). Of note, we observed that patient stratification is especially important for highly heterogeneous diseases. While some diseases (e.g. breast cancer) show few links specific to some meta-patients, more heterogeneous diseases (e.g. CNSd like schizophrenia or bipolar disorder and immune system disorders like asthma) present a majority of meta-patient specific links (*SI Appendix,* Fig. S11b). Notably, we showed that the percentage of recapitulated epidemiological interactions increased from half to 64.1% when considering the meta-patients, with a comparable or even slightly improved precision.

Even more, here we also see that many molecular similarities correspond to newly recognized comorbidities overlooked by the epidemiological network that was used as ground truth. Down Syndrome (DS), despite its clear genetic origin, showcases a wide variability in comorbid conditions (65). We found that less than 25% of its disease interactions are consistent across its meta-patients, emphasizing DS’s heterogeneity. Beyond confirming the well-established comorbidity with autism, our analysis brought to light novel associations, such as the elevated risk of leukemia in DS patients during childhood and the markedly lower incidence of various solid tumors, including breast, lung, prostate and colorectal cancers (66). Additionally, individuals with DS exhibit a higher prevalence of autoimmune disorders, with a notable sixfold increased risk of developing coeliac disease (66, 67). This susceptibility is closely linked to extensive immune system remodeling in DS, a pattern that we observe behind DS’s association with coeliac disease and lupus, another autoimmune condition that recent case studies indicate poses diagnostic challenges in DS populations (68). This example illustrates that some identified links may correspond to newly discovered comorbidities. Besides, it shows the potential of this approach to suggest new disease associations that might currently be underdiagnosed, thereby providing molecular evidence for them (*SI Appendix,* Note S14).

Indeed, incorporating the complementary epidemiological network from Hidalgo et al. based on phi-correlation allowed us to confirm that a substantial proportion of initially predicted false positives actually reflect true disease co-occurrences, yielding similar recall with markedly increased and significant precision (60%, p-value=0.0034). Further literature validation of the remaining false positives revealed robust evidence for many additional interactions, resulting in a final precision of 76.7% and suggesting that the vast majority of predicted associations correspond to true comorbidities.

As shown, the generated networks provide the first systematic translation of disease co-occurrences into molecular patterns. Previous efforts were limited by factors such as the biased knowledge of disease-associated genes for highly studied diseases or the incompleteness of the interactome, yielding small or non-significant overlaps with the epidemiology (6).

There are at least four factors that allowed us to significantly improve the explainability of known comorbidities captured with gene expression information. Firstly, the better quality and coverage of RNA-seq data, that unbiasedly increases the number and quality of features whose similarity can be compared between diseases (*SI Appendix,* Note S15). Interestingly, these features can be causally or consequentially altered in the diseases. Secondly, an improved methodology based on the existence of a significant correlation between the differential gene expression profiles across disease pairs. As a result, our disease links are robust, stable and independent of the rest of diseases, even if the disease universe changed; contrary to previous attempts based on relative molecular similarities, where disease links depended on the rest of the network (8). Thirdly, the stratification of diseases into subgroups of patients named meta-patients. Opposite to patient-centric approaches, meta-patients can be methodologically treated as phenotypes are handled in gene expression studies; thus, a significance can be associated to their altered genes, pathways and, importantly, disease interactions in the SSN (*SI Appendix,* Note S16). Lastly, this methodology allows for the characterization of negative links between diseases that can be related to inverse comorbidities, an advantage over most molecular approaches.

Still, there are a number of issues that, if addressed, could improve the quality of the results and the coverage of the comorbidity space. First, while it is possible to validate the obtained positive interactions, it is more difficult to validate the less abundant–but equally interesting–negative ones (36.63%) since they are not systematically described in large studies and are only sporadically addressed in the literature (69). Nonetheless, we detected numerous known inverse comorbidities such as: Huntington’s disease or DS and specific cancer types, Parkinson’s disease and rheumatoid arthritis, or HIV and prostate cancer (70). Therefore, a current limitation in this study is the lack of epidemiological networks entailing inverse comorbidity relationships. Also, although the epidemiological network from Hidalgo et al. provides reliable comorbidities and has been extensively used as a gold standard in the field (6, 11, 25), its comorbidities have been derived from an elderly and industrialized population. Consequently, numerous comorbidities characteristic of a younger age or different populations are not covered, some of which correspond to the observed molecular similarities (*SI Appendix,* Note S3).

Another limitation is the lack of sample information, such as age, sex, or treatments, which may drive transcriptomic differences between patients. Also, our samples belong to published studies focused on a specific disease in a given tissue (e.g. brain, liver or blood). Since we have cases and controls for each study and disease, we were able to correct for the tissue effect when generating sDEGs at the disease and meta-patient level. However, it would be optimal to have comprehensive data sets of diseases from the same tissue or an array of interesting tissues. Besides, better defined and annotated disease subgroups as well as their differential comorbidities could help us refine the definition of meta-patients and increase their power to capture their epidemiological associations.

Future perspectives include increasing sample size, so the detection power is increased (*SI Appendix,* Note S9) and sex-specific disease similarities can be extracted and compared to their epidemiologically described disease interactions (71). Expanding the analysis to noncoding regions could unveil new disease similarities potentially reflecting comorbidities (72). Furthermore, the molecular coverage of comorbidities could be improved by considering other molecular information that may underlie comorbidities within an integrative approach (73). For this, large disease cohorts comprising different kinds of omics as well as clinical information would be needed. Moreover, the identified molecular similarities could serve as a valuable guide for drug repurposing and development (74, 75).

In summary, we built disease similarity networks based on transcriptomic information that capture and meaningfully explain an unprecedented proportion of comorbidities in a significant manner. These results support the idea that comorbidities have a strong molecular component that is better captured with gene expression profiles than with other molecular sources. Actually, differential gene expression profiles portray the diseases’ altered state in a rich manner since its signal might reflect from genetic alterations to the epigenetic influence on gene expression due to internal or external factors such as treatments, contaminants or lifestyle.

This study shed light into the biological processes underlying known comorbidities, leading to a better understanding of the molecular profile and etiology of diseases and their co-occurrences. Importantly, although we showed that many of these mechanisms have been experimentally validated, our efforts propose numerous key genes and pathways that are still to be explored. Thus, we focused our discussion on some examples and provided the molecular characterization of all diseases and meta-patients at different levels of granularity (genes and pathways) within a framework that allows for the comparison of the molecular profiles for direct and inverse comorbidities in a detailed and user-friendly manner (http://disease-perception.bsc.es/rgenexcom/).

Finally, our study stresses the need to integrate the study of comorbidities and their underlying molecular similarities within a personalized medicine scope, with the aim to capture disease interactions dependent on disease subtypes or other patient-specific factors. By doing so, we aspire not only to enhance our understanding of the potential secondary conditions in specific patients but to better characterize the underlying molecular processes driving these relationships.

## Data availability

The code of the experiments is available at https://github.com/beatrizurda/Urda-Garcia_Patient_Stratification and the code of the web application can be found at https://github.com/bsc-life/rgenexcom. The data is publicly available (*SI Appendix,* Data S1); the raw data can be downloaded from GEO (https://www.ncbi.nlm.nih.gov/geo/) and the counts can be downloaded from the GREIN platform (http://www.ilincs.org/apps/grein/).

## Data Availability

The code of the experiments is available at https://github.com/beatrizurda/Urda-Garcia_Patient_Stratification and the code of the web application can be found at https://github.com/bsc-life/rgenexcom. The data is publicly available (SI Appendix, Data S1); the raw data can be downloaded from GEO (https://www.ncbi.nlm.nih.gov/geo/) and the counts can be downloaded from the GREIN platform (http://www.ilincs.org/apps/grein/).

https://github.com/bsc-life/rgenexcom

https://www.ncbi.nlm.nih.gov/geo/

http://www.ilincs.org/apps/grein/

## Acknowledgments

This work has been supported by the grants PRE2019-090454 and BES-2016-077403, both funded by MICIU/AEI/10.13039/501100011033 and by “ESF Investing in your future”; and the grants RTI2018-096653-B-I00 and PID2022-141809OB-I00, both funded by MICIU/AEI/10.13039/501100011033 and by “ERDF/EU”, by the “European Union”. The research leading to these results has received funding from the European Community’s Horizon Europe Programme under grant agreement No. 101136957 (COMMUTE). We would like to thank Davide Cirillo for their helpful comments on the work.

## Author Contributions

Beatriz Urda-García: Conceptualization, Formal Analysis, Investigation, Software, Visualization, Writing-original draft, Writing-review & editing. Jon Sánchez-Valle: Conceptualization, Supervision, Writing-review. Rosalba Lepore: Conceptualization, Supervision, Writing-review. Alfonso Valencia: Conceptualization, Funding Acquisition, Project Administration, Supervision, Writing-review & editing.

