## Supplementary Information for "Patient stratification reveals the molecular basis of disease co-occurrences"

**This PDF file includes:**

Notes S1 to S16
Figures S1 to S13
Tables S1 to S17
SI References

### Supplementary Notes

#### S1 – Assessing the significance of the network overlap with the epidemiology

We used randomizations to compute the significance of the network's overlap (recall) with the epidemiology (N=10.000). In each randomization, we rewired the ICD9 DSN preserving the degree distribution and excluding loops and computed the recall of the rewired network with the epidemiological network from Hidalgo et al. (1). Finally, we used the 10.000 generated overlaps to derive the p-value of the recall. To obtain the p-value of the precision, we followed the same approach but exchanging the networks, i.e. rewiring the epidemiological network and using the ICD9 DSN as a reference.

#### S2 – Computing the overlap of the Disease Similarity Network (DSN) with the epidemiology

We computed the overlap between the positive and negative interactions in the DSN and the ones based on medical records by Hidalgo et al. (1). To do so, we transformed the disease names in the DSN into ICD9 codes. Then, we computed the overlaps following the same methodology described in methods. In the cases in which several disease names referred to the same ICD9 code, only the interactions shared by all the diseases that correspond to that code were considered. The results of these overlaps can be found in the Table S1 and are consistent with the ones obtained by defining diseases at the ICD9 level from the beginning; the positive interactions present high and significant overlap with the epidemiology and the overlap with the negative interactions is not significant.

#### S3 – Epidemiological Network from Hidalgo et al.

The comorbidity network generated by Hidalgo et al. (1), named "Phenotypic Disease Network" (PDN), was derived from over 30 million medical records from elderly citizens (older than 65 years old) of the United States, which represented 96% of all elderly Americans.

This network connects diseases (represented as ICD9 codes) that co-occur more than expected by chance based on the significance of their relative risk, yielding undirected interactions that can be compared to molecular similarities. Other epidemiological networks provide significantly directed disease pairs (i.e. diseases that co-occur significantly and for which the directionality of that co-occurrence is also significant) (2, 3). The fact that these networks are directed and that they only include the pairs that have a significant directionality (excluding many known comorbidities) makes them less comparable to the undirected molecular similarities.

The PDN has been extensively used in the literature, specially as a gold standard of disease co-occurrences when comparing molecular similarities with comorbidities (4–6). Nonetheless, while the vast sample size ensures the robustness of the obtained interactions, some limitations of the data used to derive the network should be considered. Firstly, the data belongs to a single industrialized country. Secondly, only elderly patients were taken into account, so diseases less common at this stage of life are underrepresented (e.g. pregnancy-related diseases). Moreover, data was available only for the patients who were hospitalized at least once. As a result, this network yields robust and reliable comorbidities of its population but must miss others for which not enough information was available (e.g. comorbidities involving diseases that tend to appear at a younger age or more prevalent in other populations). The former happens for the observed relationship between breast cancer and several neoplasms (colorectal, thyroid, lung cancer or Kaposi's sarcoma), or the comorbidity between prostate and colorectal cancer, which tend to co-occur at a younger age (7). Additionally, since the PDN was built using relative risks (RR), it could overestimate comorbidities involving rare diseases and underestimate the relationships between highly prevalent diseases (1). To address this, we also assessed the overlap between our molecular networks and the PDN using phi-correlation, a complementary comorbidity measure also provided by Hidalgo et al. (1). While phi-correlation can underestimate links between rare and common diseases, it is particularly well-suited to identifying comorbidities between diseases of similar prevalence. Notably, all comorbidities captured by RR among the analyzed diseases were also found in the phi-correlation network, further reinforcing the robustness of these associations. Importantly, phi-correlation also revealed numerous comorbidities missed by RR, such as the co-occurrence of Huntington's disease with multiple sclerosis and glioblastoma, or Crohn's disease with colorectal cancer. Furthermore, we showed that the obtained molecular networks achieve consistent recall over the PDN based on phi-correlation, with considerably higher precision, indicating that a substantial proportion of the apparent false positives actually represent true comorbidities not captured by RR-based analyses.

**S4 – Manual Literature Validation of the Stratified Similarity Network**

As explained in Note S3, while the epidemiological networks from Hidalgo et al. (1) are widely used as a gold standard for disease co-occurrence, they present certain limitations. These networks are based on data from a single industrialized country, include only elderly individuals, and consider only patients who were hospitalized at least once. As a result, although they provide robust and well-validated comorbidities, they may fail to capture true disease co-occurrences that are more prevalent in other populations or those involving younger individuals, rare diseases, or conditions underrepresented in hospital settings.

To refine the evaluation of our molecular interactions, we first incorporated the complementary epidemiological network from Hidalgo et al., based on phi-correlation. This measure is particularly effective at capturing comorbidities between diseases of similar prevalence, which relative risk (RR) may underestimate. Including the phi-based network in our validation notably increased the precision of our method (60%, p-value = 0.0034) while maintaining consistent recall (63.1%, p-value = 0.0025), indicating that many of the initially predicted false positives using the RR-based network are in fact supported by epidemiological data (Table S8).

Nevertheless, as expected from the mentioned limitations of these networks, some interactions remained labeled as false positives even after including the phi-correlation network. To evaluate these remaining cases, we performed a manual literature validation of the top and bottom 100 false positive interactions in the SSN, ranked by correlation strength. For each interaction, we searched PubMed and PubTator3, which retrieves publications containing specific relations between two entities, such as diseases, accounting for name variants and standardized identifiers. We also used Semantic Scholar, a literature search engine with flexible NLP-based querying, useful for identifying relevant biomedical publications, especially in cases where terminology may vary across studies. Only associations supported by strong epidemiological evidence–such as large cohort studies, meta-analyses, or consistent findings across multiple studies–were considered validated. Interactions for which evidence was ambiguous or inconclusive were marked as uncertain and excluded from the final count of validated positives.

This manual curation revealed that approximately 30% of these remaining interactions were indeed supported by the literature. When combined with the 30% already captured by the phi-correlation network, we found that 60% of the false positives identified using the RR-based network actually correspond to true disease co-occurrences. Incorporating this correction increased the final precision of our method from 43.06% to 76.65%, demonstrating that the vast majority of the obtained disease associations correspond to true comorbidities, even if initially missed by standard epidemiological networks.

We further assessed the performance of our method by validating all SSN interactions between neoplasms, one of the disease category pairs with the lowest reported precision from the reference epidemiological network. While the networks from Hidalgo et al. capture numerous established associations between cancers–such as the co-occurrence of lung cancer with liver and colorectal cancer, chronic lymphocytic leukemia, Kaposi's sarcoma and glioblastoma–they fail to detect a considerable number of other well-supported links. This underdetection may stem from the cohort's demographic bias, focused on elderly hospitalized individuals, which limits the detection of comorbidities that tend to appear earlier in life or in other populations. For instance, this is the case for the co-occurrence of breast cancer with colorectal, thyroid, lung cancer, and Kaposi's sarcoma, or the comorbidity between prostate and colorectal cancer–interactions frequently reported in younger cohorts (7). Notably, 69.7% of the neoplasm–neoplasm interactions missed by the reference network were found to be supported by large-scale epidemiological studies with longer follow-up periods, suggesting that the lower precision observed for this category largely reflects true comorbidities not captured by the standard reference (Table S17). Interestingly, the above-mentioned breast cancer interactions have also been predicted by computational methods that leverage drug-indication and drug-target inference for comorbidity detection, supporting their functional relevance and interest for drug repurposing strategies (8).

This analysis underscores the importance of integrating complementary validation strategies–including complementary epidemiological metrics and manual literature curation–for a more comprehensive and accurate evaluation of disease associations. It also highlights the strength of our method in uncovering previously undetected comorbidities, such as those involving rare or underrepresented diseases that may be overlooked in traditional epidemiological studies due to population biases or data limitations.

Notes S14 and S15 discuss several of these newly identified disease co-occurrences, proposing novel mechanistic explanations for them and suggesting additional disease co-occurrences with molecular evidence.

Notably, we encountered several interactions for which validation was not possible due to the lack of published studies, despite strong molecular signals. For example, familial dysautonomia is a rare neurodevelopmental disorder with a higher prevalence among individuals of Ashkenazi Jewish descent and fewer than 600 described cases in the medical literature (9). With an average life expectancy in the third decade, it remains poorly studied, particularly in terms of comorbidities. Even so, we were able to detect multiple known disease co-occurrences involving familial dysautonomia, including associations with autism, bipolar disorder, Huntington's disease, lupus, rheumatoid arthritis, muscular dystrophies, Crohn's disease, coeliac disease, and ulcers. Furthermore, we identified several significant links with neoplasms, including the established co-occurrence with glioblastoma and various types of polyps. Although previous studies suggest that patients with familial dysautonomia may be at increased risk for developing tumors (10, 11), and the causal gene has been implicated in tumorigenesis (11), the lack of epidemiological research prevented us from confirming whether other observed associations–such as those with lung, breast, liver, prostate cancer, and Kaposi's sarcoma–reflect true comorbidities.

Additional potentially valid but currently unconfirmed associations due to limited available literature include links between Alagille syndrome and Parkinson's disease, Huntington's disease and ischemia, coeliac disease and glioblastoma, autism and glioblastoma, multiple sclerosis and Parkinson's diseases, Lyme disease and chronic obstructive pulmonary disease (COPD), colorectal cancer and HIV, or breast cancer with Alagille syndrome or muscular dystrophies, among others (Table S16). These findings suggest that the current precision estimate of 76.65% may represent a conservative lower bound, as several strong molecular associations could not be validated due to the absence of epidemiological data. Future studies may help confirm these associations and further refine the precision estimate.

### S5 – Network backbone

To aid the visualization of the networks, we obtained the network backbone of the DSN and the SSN by applying the method from Simas et al. (12) using the metric and ultra-metric closures. We generated the network backbone separately for the positive and negative interactions, considering the edge's weight as 1 - |Spearman's correlation| + ε, where $\varepsilon = 10^{-6}$. Then, we defined the network backbone of the entire network as the union of the positive and negative subnetwork's backbones. Table S3 shows the percentage of interactions in the positive, negative and entire DSN and SSN that are kept in the network backbones. When considering the metric closure, 96% of the DSN are kept, indicating a low level of redundancy. However, 45% of the interactions are kept for the SSN, which exhibits higher redundancy in its positive than negative interactions. As expected, the more restrictive ultra-metric closure yields smaller backbones of 13.4 and 3.7% for the DSN and SSN, respectively.

### S6 – Positive and negative interactions in the DSN

Positive interactions in the Disease Similarity Network (DSN) are defined as significant positive Spearman's correlations between the differential gene expression profiles of two diseases (Methods). Specifically, these profiles correspond to vectors of log fold changes in gene expression relative to controls for each disease. Therefore, a positive correlation indicates that genes upregulated in one disease also tend to be upregulated in the other, and likewise for downregulated genes.

Biologically, such correlations suggest shared molecular alterations, including activation or repression of common pathways, similar regulatory responses, or overlapping cellular processes. While a positive interaction does not necessarily imply a direct functional dependency, it reflects convergent transcriptomic programs that may arise from shared etiology, pathophysiological mechanisms, or compensatory responses.

Throughout the manuscript, we discuss several use cases in which positively linked diseases share the significant dysregulation of genes and pathways previously shown to underlie their comorbidity or pathology, as well as molecular mechanisms that may represent novel insights into these relationships. Examples include the Parkinson's disease and asthma co-occurrence (Note S14), the Parkinson's–glioblastoma link (Note S15), the comorbidities of Down syndrome (Discussion), or the stratified breast cancer links (Discussion and Note S13).

In contrast, negative interactions correspond to significant negative correlations between the differential expression profiles of two diseases. These imply that genes upregulated in one disease tend to be downregulated in the other, and vice versa. Such inverse patterns may reflect opposing molecular alterations, including inverse regulation of pathways or divergent physiological states. For instance, we identified robust negative interactions between Huntington's disease (HD) and several cancer types, including liver, lung, and breast cancer. In these cases, 85% of the pathways significantly altered in both diseases are dysregulated in opposite directions (e.g. overexpressed in HD and underexpressed in cancers). Notably, HD and breast cancer exhibit opposite significant dysregulation in over 600 genes and pathways, demonstrating that these interactions reflect meaningful biological differences at multiple levels of molecular resolution (see Note S8 for a detailed discussion of this use case).

**S7 – Sample size impact on network structure**

We performed saturation analyses to assess whether the relationship between the number of significantly differentially expressed genes (sDEGs) and sample size is an artifact of smaller sample sizes (Methods). We observed that, as expected in gene expression studies, this correlation exists (13) and remains consistent across increasing sample size thresholds even after removing two-thirds of the diseases in the dataset (Fig. S13a).

We also examined the moderate association between node degree and sample size. Figure S3b shows that many diseases with fewer than 50 samples exhibit node degrees comparable to those of diseases with larger sample sizes. This moderate trend aligns with biological expectations, where larger sample sizes may provide higher statistical power to detect interactions. Importantly, this trend also persists after saturation analysis (Fig. S13b), confirming that while smaller sample sizes are generally associated with fewer sDEGs and interactions, they do not distort the expected network structure. For a detailed discussion on why sample size differences do not introduce biases in downstream results, see Note S9.

**S8 – Opposing Molecular Signatures Between Huntington's Disease and Cancer**

To explore the molecular basis of the negative correlations observed between Huntington's disease (HD) and several cancer types, we examined significantly altered pathways shared between HD and liver, lung, and breast cancer, as well as chronic lymphocytic leukemia (CLL). We found that, on average, 85% of the significantly dysregulated pathways shared between HD and these cancers exhibit opposite regulation. Specifically, 71% in liver cancer, 77% in lung cancer, 92% in breast cancer, and 100% in CLL. Indeed, HD shares the opposite regulation of numerous significantly differentially expressed genes and pathways, with breast cancer exhibiting over 1000 genes and 600 pathways. Many of these processes are consistently altered in the same direction across multiple cancer types, suggesting a broad and robust divergence in molecular profiles.

Among the most consistent patterns, *SOD2* (encoding mitochondrial superoxide dismutase 2) is overexpressed in HD, likely as a compensatory response to oxidative stress and mitochondrial dysfunction (14–16). In contrast, *SOD2* is underexpressed in all four cancer types, where reduced antioxidant activity is associated with increased oxidative damage and tumor progression (17). Likewise, the pathway positive regulation of reactive oxygen species biosynthetic process is overexpressed in HD and underexpressed in these cancers.

Immune-related processes also show contrasting patterns. Pathways such as cytokine production involved in immune response, as well as interleukin-1β, IL-1, and IL-8 signaling and production are overexpressed in HD but underexpressed in at least three cancer types, in line with HD's neuroinflammatory profile. In HD, chronic activation of pro-inflammatory chemokines contributes to neuronal damage, whereas cancers often suppress immune signaling and cytotoxic immune cell infiltration to promote immune evasion (18, 19).

Apoptosis regulation also reveals contrasting signatures. *TP53BP2*, a gene that promotes p53-mediated apoptosis, is overexpressed in HD, where it may contribute to neuronal apoptosis, but underexpressed in all the cancers, consistent with apoptotic evasion in tumorigenesis.

Additionally, several genes encoding subunits of mitochondrial complex I, including *NDUFS1*, *NDUFV1, NDUFS8, NDUFA9*, and *NDUFB6*, are overexpressed in HD, likely reflecting compensatory mitochondrial response. These same genes are underexpressed across cancers, a pattern generally observed especially for those relying on aerobic glycolysis (Warburg effect) rather than mitochondrial respiration to support rapid proliferation (20, 21).

These results support the existence of robust opposite molecular profiles underlying the observed inverse comorbidities between HD and multiple cancer types, spanning core cellular processes such as oxidative stress response, immune signaling, mitochondrial metabolism, and apoptosis.

**S9 – Sample size effect on the recall and precision**

To ensure that the obtained results are not affected by including diseases with smaller sample sizes, we performed saturation analyses on precision and recall. Specifically, we computed the recall and precision of the Disease Similarity Network (DSN) over the epidemiological interactions in Hidalgo et al. (1) for increasing sample size thresholds following the procedure described in Methods. These analyses demonstrated that both precision and recall remain stable even when progressively removing diseases with lower sample sizes (Fig. S13c-d). Notably, our conclusions remain consistent and statistically significant even when retaining only 30% of the interactions, confirming the robustness of the approach to sample size constraints.

To further assess the impact of sample size on disease co-occurrence detection performance, we computed the F1-score (the harmonic mean of precision and recall) for each disease in the DSN. We found no correlation between F1-score and sample size (Spearman's R = 0.263, p-value = 0.101), indicating that co-occurrences of diseases with both smaller and larger sample sizes are captured at a similar level. A more detailed analysis revealed that while recall is moderately correlated with sample size (Spearman's R = 0.33, p-value = 0.035), precision is not affected (p-value = 0.08). This suggests that smaller sample sizes may modestly limit interaction detection (leading to lower recall and node degree), but they do not increase false positives, as precision remains stable[1]. Importantly, this confirms that including diseases with smaller sample sizes does not introduce systematic bias or degrade overall performance.

Several diseases with small sample sizes further illustrate this robustness. Kaposi's sarcoma (n=8), a rare vascular tumor, achieves above-average recall (63.3%) and precision (73.1%), demonstrating that its disease co-occurrences are well captured despite its limited data. The DSN identifies 19 validated disease co-occurrences, plus its recently described association with breast cancer, reinforcing the biological relevance of our findings. Similarly, familial dysautonomia (n=8), a rare genetic disorder, also presents above-average recall and precision, with the DSN capturing 14 significant disease co-occurrences supported by molecular

[1] Consistent results were obtained when comparing the DSN with the epidemiological network based on relative risks (RR). As expected, disease sample size significantly correlates with recall (Spearman's correlation = 0.33, p-value = 0.039); however, no correlation was observed with precision (p-value = 0.81). The lack of an effect on precision can be explained by the fact that only disease pairs with significant correlations after multiple testing are selected, thereby reducing the likelihood of false positives.

similarities. Rheumatoid arthritis (n=14) exhibits slightly lower recall (3.5 percentage points below average), yet the DSN identifies 9 interactions, all of which have been previously reported as comorbid (precision=100%). These examples illustrate that even rare diseases with lower sample sizes retain robust and biologically meaningful interactions, reinforcing that larger sample sizes are not a prerequisite for detecting disease co-occurrences. Since rare diseases are generally less characterized, this property is especially valuable, as it may enable the identification and molecular exploration of co-occurrences that may remain inaccessible to molecular approaches relying on prior knowledge.

Additionally, the definition of meta-patients showed that actually more disease interactions can be recapitulated by splitting the diseases into subgroups, which necessarily reduces sample size, due to the existence of transcriptomic heterogeneity. This effect was observed for all analyzed diseases (Fig. S11a). In fact, some diseases present a two-fold increase in the number of disease links with the definition of meta-patients (e.g. muscular dystrophy or autism). Notably, the definition of meta-patients leads to a considerable increase in recall while maintaining precision, further underscoring the robustness of the method in capturing biologically relevant interactions across diseases and sample sizes.

Taken together, these findings confirm that while larger sample sizes may increase power to detect additional interactions, our approach remains effective in capturing meaningful and biologically relevant co-occurrences even for diseases with limited sample sizes, such as rare diseases. Our methodological approach is robust to sample size variation, ensuring reliable conclusions across diverse sample size thresholds. Nonetheless, larger sample sizes are needed to increase detection power, allowing for the stratification of this study based on clinically relevant features, such as sex, age, or ancestry.

### S10 – Within- and Between-Category Interactions in the Disease Similarity Network (DSN)

We evaluated whether the DSN tends to connect diseases from the same ICD9 category by computing the network's assortativity with respect to disease categories. The resulting near-zero assortativity value confirms that the DSN does not preferentially link diseases from the same category. This aligns with the reference epidemiological network, which also exhibits a neutral tendency for within-category links (Table S5).

To further assess whether the DSN primarily reflects broad disease classifications or captures more specific molecular similarities, we analyzed the distribution of interactions within and between ICD9 disease categories. Only 25.2% of the positive interactions in the DSN connect diseases within the same category, indicating that the majority of links occur between diseases from different categories. To test whether within-category interactions tend to show stronger transcriptional similarity, we compared the distribution of Spearman's correlation values for within- and between-category disease pairs. A Wilcoxon rank-sum test revealed no significant difference between the two distributions (p-value = 0.44), suggesting that both types of interactions are similarly strong.

Together, these results indicate that the DSN captures molecular relationships that transcend conventional disease classifications. We highlighted several between-category disease links throughout the manuscript (e.g. Down Syndrome or breast cancer interactions). Additionally, Note S14 shows how other underlying factors–such as chronic inflammation–may drive both the observed molecular similarities and the newly identified comorbidities between Parkinson's disease (nervous system) and diseases from other categories, including asthma (respiratory), coeliac disease (digestive system), lupus (musculoskeletal system and connective tissue), and potentially other conditions characterized by chronic inflammation.

**S11 – Comparison of the Disease Similarity Network (DSN) with the PPI and microarray-based networks**

We compared the networks that present the largest significant overlap with the epidemiology (PPI and microarray-based networks) with the DSN (Table S11–12). The network derived from PPIs (4) and the DSN share 19 ICD9 codes and only 6 out of the 20 interactions present for these diseases in the former are found in the later (there is no significant overlap between them). Between these common diseases, 29 epidemiologically known comorbidities are connected only in the DSN whereas 10 are unique to the other network (Fig. 5b). The entire DSN provides information for 22 new ICD9 codes and uniquely captures 149 disease links described in the epidemiology.

On the other hand, the microarrays' network (22) contains 92 ICD9 codes, where 27 of them are analyzed in the DSN. We computed the overlap of both networks over the common set of ICD9 codes, yielding a significant overlap (p-value = 0.027) of 47.02% of the microarrays' network. Specifically, positive interactions have a significant overlap (p-value = 0.002) of 62.22% whereas the overlap of the negative interactions is not significant (p-value = 0.624) (Table S12). Among these common diseases, the DSN yielded 42 new positive interactions that are described in the epidemiological network by Hidalgo et al. (1) (e.g. Crohn's disease and ulcerative colitis) (Fig. 5c). Additionally, the DSN provides information for 14 new ICD9 codes and captures 141 new interactions that match known comorbidities.

**S12 – Common overexpression of immune system pathways in comorbidities**

Almost all the detected epidemiological interactions (EIs) in the DSN share the significant overexpression of at least one–and a mean of 21.2–immune system pathways. The top list of commonly overexpressed immune system pathways in EIs can be found in table S13 (e.g. interferon signaling, antigen presentation and multiple interleukins signaling). Actually, the immune system is the Reactome pathway category (23) that underlies the highest number of observed EIs, pointing towards its key role in the development of comorbidities. Indeed, previous efforts have shown that diseases often share the alteration of immune system genes or pathways. Li et al. extracted disease associated genes from literature mining, generated their enriched pathways and built a disease-disease network connecting diseases based on their shared pathways (24). They found that the immune system pathways were the ones underlying the highest number of diseases and tended to be simultaneously altered in multiple ones. Barrenas et al. followed a different approach based on GWAs and concluded that most genes associated with more than one disease were inflammatory (25). They also observed an overrepresentation of inflammatory genes in the disease-associated genes of complex diseases, where several of the top enriched pathways match our top immune system pathways in EIs (e.g. cytokines, antigen processing and presentation) (Table S13). Similarly, Suthram et al. used PPIs and gene expression data to show that most of the disease modules commonly altered in over 50 diseases are mainly enriched in immune system and DNA repair processes (26). Put in context, these results suggest that most diseases that are indeed comorbid share the overexpression of a variety of immune processes (e.g. interferon alpha or beta signaling, antigen presentation involving MHC I, or interleukin 4 and 13 underlie 51, 51 and 41 EIs respectively). Thus, the overexpression of these pathways could be at the essence of numerous comorbidities.

**S13 – Potential molecular mechanisms behind the specific links of breast cancer meta-patients**

Autism and bipolar disorder are positively linked to ER- and TN breast cancer meta-patients. Autism shares the overexpression of 68 overexpressed and 27 underexpressed pathways from Reactome (23), KEGG (27) and GO (28) with ER- and TN meta-patients whereas bipolar disorder presents more than 100 pathways in common with them. Autism exhibits an extraordinarily high heritability, an extensive overlap in risk genes and pathways with cancer (29) and a significant overlap of significantly altered pathways with several cancer types (30). Crawley et al. indicate that both phenotypes share the alteration of a myriad of biological functions, many of which we find commonly altered between autism and these breast cancer meta-patients (e.g. chromatin remodeling, DNA repair and maintenance, cell proliferation and apoptosis or p53 and PTK7 activity) (29).

Multiple sclerosis (MS), a complex inflammatory disease characterized by CNS lesions (31), is negatively linked to ER+ and ER- breast cancer meta-patients. More than 80 pathways are significantly differentially altered in opposite directions in these phenotypes. Actually, most of them are immune system pathways (inflammatory response, lymphocytes activation and differentiation, macrophages activation or the production of several interleukins) that are significantly overexpressed in MS and underexpressed in these meta-patients, matching the diseases' pathogenesis (31, 32) and potentially underlying their negative relationship. This opposite pattern of expression is also observed for other processes, such as apoptosis, insulin-like growth factor (IGF) receptor signaling or toll-like receptor (TLR) signaling. Interestingly, the IGF system has been shown to be involved in both diseases; IGF-1 has been considered a therapeutic agent for MS due to its neuroprotective and myelinogenic capacity (33) whereas many therapeutic strategies for breast cancer have been based on targeting the IGF-1 receptor, involved in cell growth and linked to therapy resistance (34). Similarly, TLRs proinflammatory activity has been previously linked to MS, where low doses of TLR 4 antagonists are being tested in clinical trials for MS and other autoimmune disorders (35, 36). On the other hand, TLRs have been shown to be underexpressed in breast cancer and related to the disease occurrence, subtype and development (37).

### S14 – Chronic inflammation and neurodegeneration: molecular insights into the asthma–Parkinson's disease Link

Many of the molecular similarities identified between diseases correspond to newly detected comorbidities, which were missed by Hidalgo et al. (1) (Note S3–4). One particularly interesting case is the association between asthma and Parkinson's disease (PD). Our analysis reveals a robust molecular association between asthma and PD (p-value = 8.14e-95).

Very recently, a nationwide retrospective cohort study has confirmed this association, showing that individuals with asthma are at a higher risk of developing PD, even after adjusting for confounding factors such as lifestyle (38). An independent nationwide cross-sectional retrospective study further supports this finding, revealing a dose-dependent relationship, where PD risk increases with asthma severity (39). While clinical hypotheses have been proposed to explain this connection, to our knowledge, no molecular studies have systematically investigated it to date.

Our findings reveal that asthma and PD share the significant alteration of over 250 genes, highlighting the shared overexpression of key immune system pathways, including interferon alpha and beta signaling, leukocyte-mediated immunity, and T-cell regulation. These processes play a central role in neuroinflammation and epithelial barrier integrity, suggesting a molecular basis for the observed epidemiological link.

These findings provide molecular evidence supporting the epithelial barrier hypothesis and the lung-brain axis theory as potential mechanisms linking chronic inflammation in asthma to neurodegeneration in PD. Persistent airway inflammation can disrupt epithelial barriers in the lungs, leading to systemic immune activation and increased circulating proinflammatory cytokines (40). These cytokines may then enter the central nervous system via the blood-brain barrier, triggering neuroinflammation. Peripheral immune cell penetration leads to microglial activation in the brain, particularly present in the substantia nigra (41), a key region affected in PD. Once activated, microglia release reactive oxygen species (ROS) and neurotoxic cytokines, which can promote dopaminergic neuronal degeneration, a hallmark of PD.

Furthermore, both diseases share the overexpression of wound healing and angiogenesis pathways, suggesting tissue remodeling in response to chronic inflammation (42). While these processes remain largely uncharacterized in PD, emerging evidence suggests new vessel formation in PD that may lack key restrictive properties of the blood brain barrier (43). This could potentially facilitate the entry of proinflammatory cytokines into the central nervous system, further driving neurodegeneration and exacerbating PD pathology.

Neuroinflammation is increasingly recognized as both a risk factor and a potential contributor to PD pathogenesis (38, 44, 45). The strong molecular overlap between these diseases provides a plausible mechanistic explanation for their epidemiological association and suggests that targeting immune pathways may offer new therapeutic opportunities for patients and individuals at risk. Indeed, adequately managing inflammatory processes may not only improve asthma symptoms but also reduce the risk of potential secondary conditions, such as PD. Notably, Bower et al. demonstrated that asthmatic patients who used anti-inflammatory drugs had a lower risk of developing PD, further supporting the proposed mechanism (45).

Given the strong connection between asthma and PD, we sought to determine whether other diseases characterized by chronic inflammation also co-occur with PD. Indeed, our analysis revealed significant molecular associations between PD and other chronic inflammatory diseases, including coeliac disease, lupus, and Crohn’s disease, all of which exhibit marked overexpression of immune system processes shared with PD. Interestingly, these comorbidities were missed by the epidemiological network based on relative risks Hidalgo et al. (1) but were confirmed when incorporating the epidemiological network based on phi-correlation (coeliac disease) or through large-scale studies specifically examining these associations (Table S16). These findings further support the role of chronic inflammation as a risk factor for PD development, with diseases such as asthma, coeliac disease, lupus, and Crohn's disease–and likely others–potentially contributing to or exacerbating neurodegeneration.

Future epidemiological studies may identify additional inflammatory diseases that increase PD risk, further refining our understanding of inflammation-driven neurodegeneration. Ultimately, managing inflammation is not only critical for autoimmune diseases but may also be key to neurodegenerative disease prevention and treatment, reinforcing the need for further research into immune modulation as a therapeutic strategy in PD.

This use case highlights the power of our approach to capture known disease co-occurrences and to identify novel disease pairs with molecular evidence, providing mechanistic explanations for them that could inform targeted therapeutic strategies. By bridging molecular and epidemiological data, it offers deep insights into systemic disease relationships, suggesting potentially unrecognized disease associations, and contributing to disease characterization.

**S15 – Molecular insights into the Parkinson's disease–Glioblastoma link**

While Parkinson's disease (PD) has been widely reported to be associated with a decreased risk of several cancers, such as liver cancer, recent studies suggest that PD patients have an increased risk of brain cancers, including glioblastoma (46–49).

In line with these findings, our analysis uncovers a strong molecular association between PD and glioblastoma (Spearman's correlation = 0.49, p-value = 0). These diseases share 79 significantly overexpressed and 30 underexpressed Reactome pathways, along with over 2,500 differentially expressed genes altered in the same direction, suggesting common underlying molecular mechanisms. Among these genes, a key molecular link is the *PARK2* gene, whose mutations are the most common cause of early-onset PD and have been recently implicated in glioblastoma development due to their tumor suppressor role (50). Moreover, both diseases exhibit reduced dopamine secretion and transport. PD is characterized by the progressive degeneration of dopaminergic neurons in the substantia nigra, leading to a substantial decrease in dopamine levels. Although the role of dopamine in glioblastoma is complex, studies suggest that dopamine may exert anti-tumor effects, to the extent that therapeutic strategies targeting dopamine receptors are being considered (51). This raises the possibility that dopamine deficiency in PD may influence glioblastoma risk, warranting further investigation.

Additional biological processes that may underlie this interaction include immune system activation, collagen and extracellular matrix remodeling (e.g. integrins and proteoglycans), increased angiogenesis, signaling processes (e.g. regulated by *TP53*, *RUNX3*, *PTK2*, *SMAD2-4*, or the insulin-like growth factor), and epigenetic regulation (e.g. via PRC2 (52)). A detailed examination of these shared molecular mechanisms may offer new insights into the link between PD and glioblastoma, potentially leading to novel therapeutic strategies.

This case illustrates how low-prevalence disease co-occurrences may be difficult to detect in epidemiological studies, yet molecular evidence can effectively highlight plausible associations, providing a basis for exploring their underlying mechanisms.

Importantly, our approach goes beyond confirming known trends by uncovering new mechanistic insights and directional disease associations (see other use cases in Note S13, Note S14, and the Discussion). A key factor enabling the detection of robust and significant disease similarities–even for pairs missed by previous epidemiological studies–is the broader coverage of RNA-seq data. Unlike other approaches that may be limited by biased knowledge on disease-associated genes for highly studies diseases or the incompleteness of the interactome (e.g. PPI networks), RNA-seq provides quantitative gene expression profiles across all genes for all the analyzed diseases.

Moreover, transcriptomic data can capture coordinated changes in gene regulation that may underlie disease co-occurrences via mechanisms not yet characterized (as illustrated for the PD–glioblastoma example). In contrast, other molecular data types such as PPI networks often lack regulatory information, including the directionality in which genes are altered. Gene expression data allows us to quantify both the magnitude and direction of gene expression alterations–whether genes are over- or underexpressed–enabling the detection of both positive and negative disease associations. By leveraging this feature, we can distinguish between direct comorbidities (pairs that tend to co-occur more frequently than expected by chance) and inverse comorbidities (those that co-occur less). For instance, even when similar genes are altered in PD–liver cancer and PD–glioblastoma, taking quantitative and directed gene expression measures into account allowed us to correctly identify an inverse relationship with liver cancer and a direct comorbidity with glioblastoma. This differentiation enables us to propose mechanistic hypotheses for disease associations that may appear similar on the surface but differ fundamentally in their biological underpinnings and their clinical implications.

**S16 – Comparison with Sánchez-Valle et al.**

A previous work from Sánchez-Valle et al. (22) derived molecular similarities between diseases and patients from microarray data, finding a small but significant overlap of 16% with the epidemiological network from Hidalgo et al. (1). On the contrary, this work uses RNA-seq data, which is currently replacing microarrays due to its improved sensitivity, reproducibility and detection's dynamic range. The fact that microarrays have lower quality and are in disuse, while RNA-seq has proved superior quality and has a growing number of publicly available data (Fig. S1) is an important factor that ensures the usefulness and scalability of the present work.

Importantly, this work presents a different approach with improved methodology that allows to significantly capture a high percentage of comorbidities for the first time (49% vs. 16%). In Sanchez-Valle et al. (22), similarities are extracted at the patient level by computing Fisher's exact test based on the top 500 over and underexpressed genes. Then, Relative Molecular Similarities are calculated to derive disease-disease similarities from the similarities at the patient level. Alternatively, here we have computed Spearman's correlations between the actual diseases' differential expression profiles. Therefore, no cut-offs are needed and the addition of new diseases does not change the existing disease relationships. In other words, now the disease links are more robust, stable and independent of the rest of diseases.

Moreover, Sánchez-Valle et al. (22) generated disease subgroups by clustering patients based on their differential gene expression profile and used Relative Molecular Similarities to derive their links. This approach hindered the obtention of significantly altered genes and pathways at the disease subgroup level. On the contrary, in this work, we have clustered patients based on their gene expression profiles and considered them as phenotypes, named meta-patients. This allows us to characterize meta-patients based on their significantly differentially expressed genes and pathways and to robustly extract and explain their molecular similarities with other meta-patients or diseases. Moreover, we are the first to consider and compute the overlap with the epidemiology at the level of disease subgroups, achieving a considerably higher recall of 64% with a sustained precision. Thus, meta-patients are the first step towards the robust stratification of the study of comorbidities at the level of disease subgroups, which the results strongly advocate for. In conclusion, this new approach based on more updated and higher quality data shows, for the first time, that a strong correspondence exists between disease co-occurrences and molecular similarities at the disease and subgroup level (significant recalls of 49% and 64% respectively).

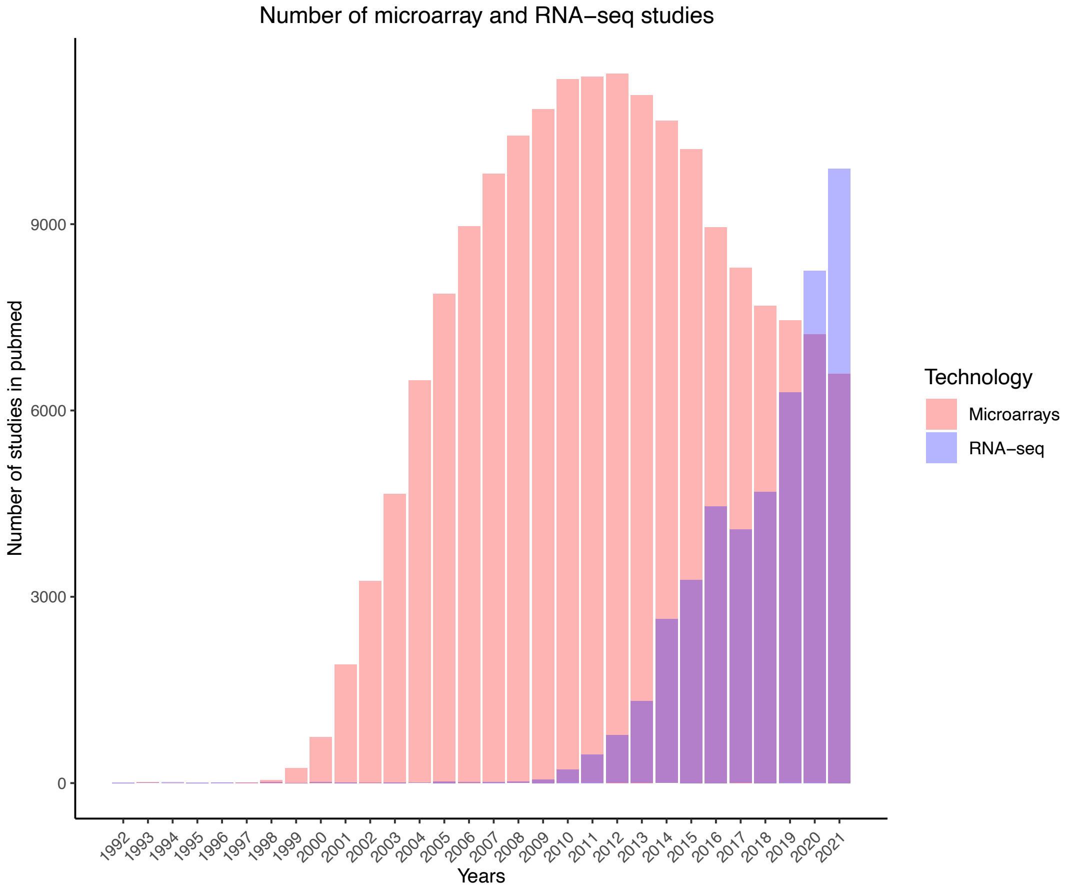


**Fig. S1. Evolution of the number of microarray and RNA-seq studies in PubMed.** Number of studies in PubMed using microarray and RNA-seq technologies from 1992 to 2021. The data was downloaded directly from PubMed on the 21st of January 2022.

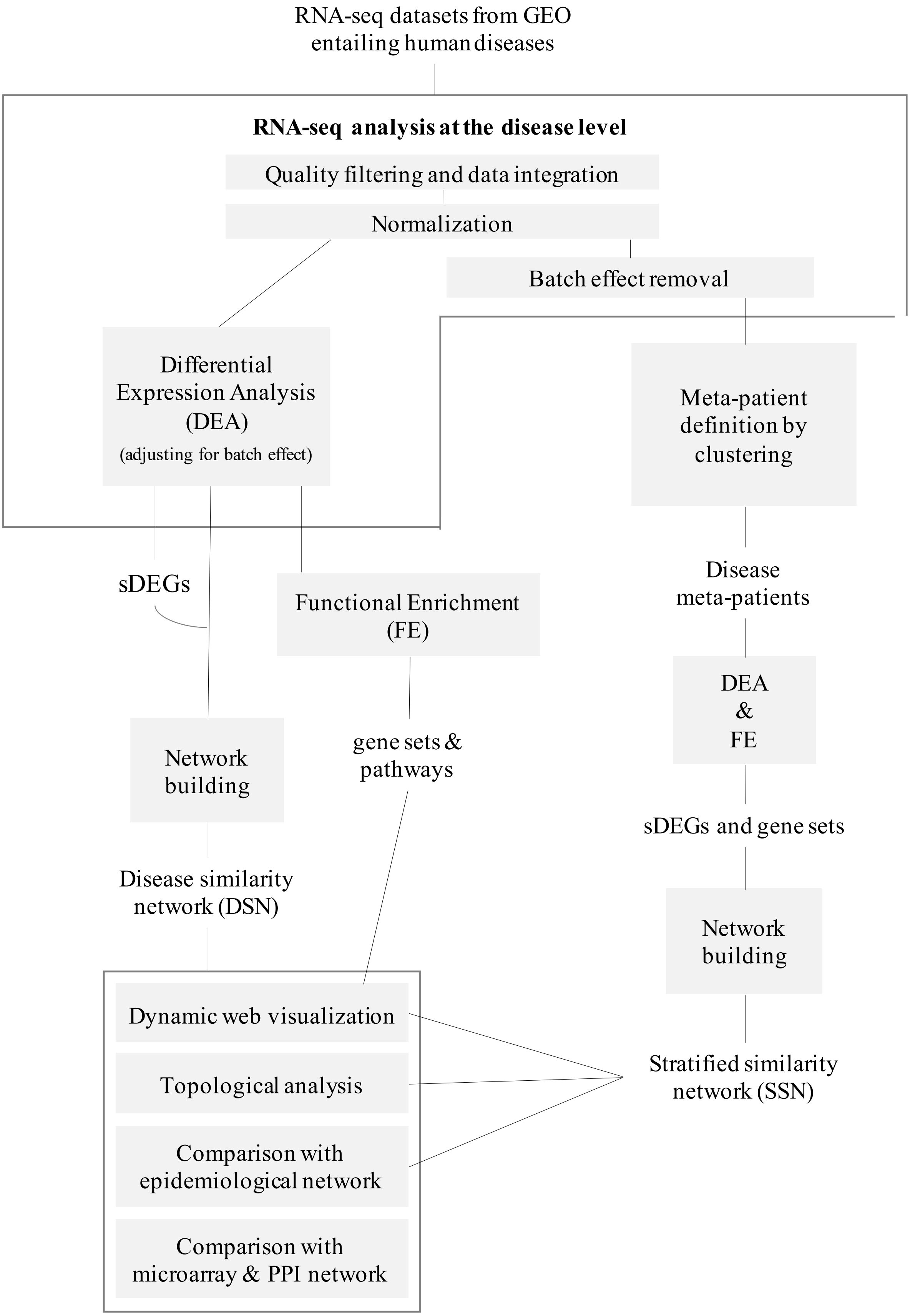
RNA-seq datasets from GEO entailing human diseases
RNA-seq analysis at the disease level
Quality filtering and data integration
Normalization
Batch effect removal
Differential Expression Analysis (DEA)
(adjusting for batch effect)
Meta-patient definition by clustering
sDEGs
Functional Enrichment (FE)
Disease meta-patients
DEA & FE
Network building
gene sets & pathways
sDEGs and gene sets
Disease similarity network (DSN)
Network building
Dynamic web visualization
Topological analysis
Comparison with epidemiological network
Comparison with microarray & PPI network
Stratified similarity network (SSN)

**Fig. S2. Analysis pipeline schema.** Schematic representation of the workflow employed in this study. Initial steps involved the collection of RNA-seq datasets from the Gene Expression Omnibus (GEO) featuring human diseases. Subsequently, RNA-seq analysis was conducted at the disease level, encompassing quality filtering, data integration and normalization. Differential expression analyses (DEA) were then applied to obtain significantly differentially expressed genes (sDEGs) for each disease. Functional enrichment (FE) was performed to identify the significantly differentially expressed gene sets and pathways for each disease. Additionally, we built a Disease Similarity Network (DSN) based on the similarity between the differential gene expression profiles of diseases. Besides, we obtained disease meta-patients by applying clustering algorithms to the normalized and batch effect corrected counts for each disease. DEA and FE were then performed on the meta-patients, and a Stratified Similarity Network (SSN) was created, incorporating the meta-patients into the DSN. The topological properties of the DSN and the SSN were analyzed, and both networks were compared with the epidemiological network from Hidalgo et al. (1, 53). The DSN was further compared with other disease-disease networks based on molecular information, including microarray (22) and protein-protein interaction (4) data. Finally, a functional web application was developed for easy inspection of the networks and their underlying molecular mechanisms.

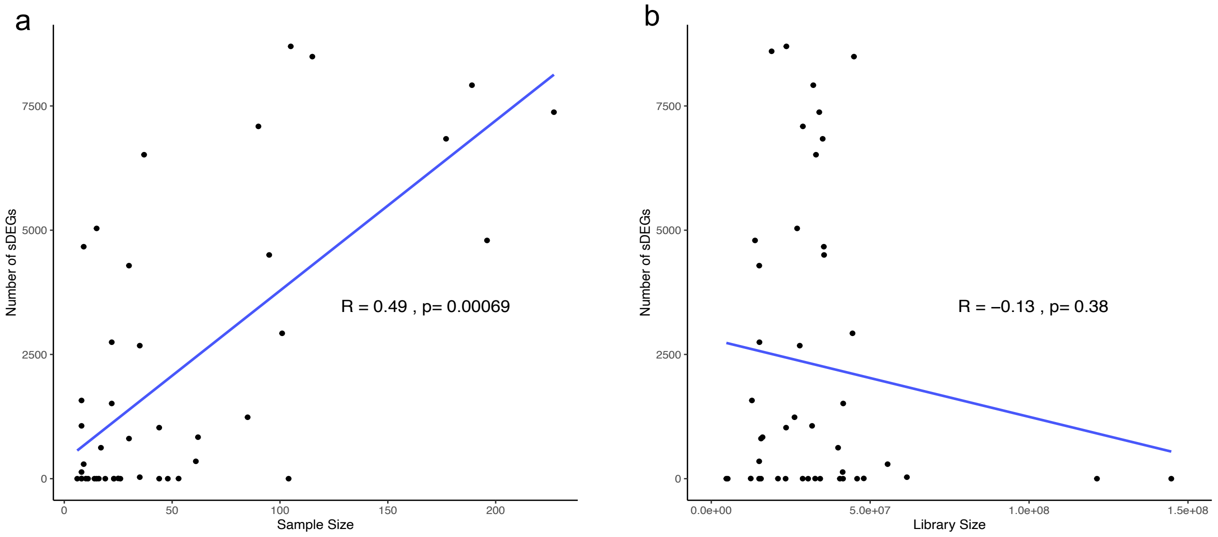


**Fig. S3. Correlation between the number of significantly differentially expressed genes (sDEGs) with sample size and library size**. (**a**) Spearman's correlation between the number of sDEGs and sample size in our disease set. (**b**) Spearman's correlation between the number of sDEGs and the average library size in our disease set. The library size, defined as the average number of sequencing reads per sample, serves as a metric for sequencing depth.

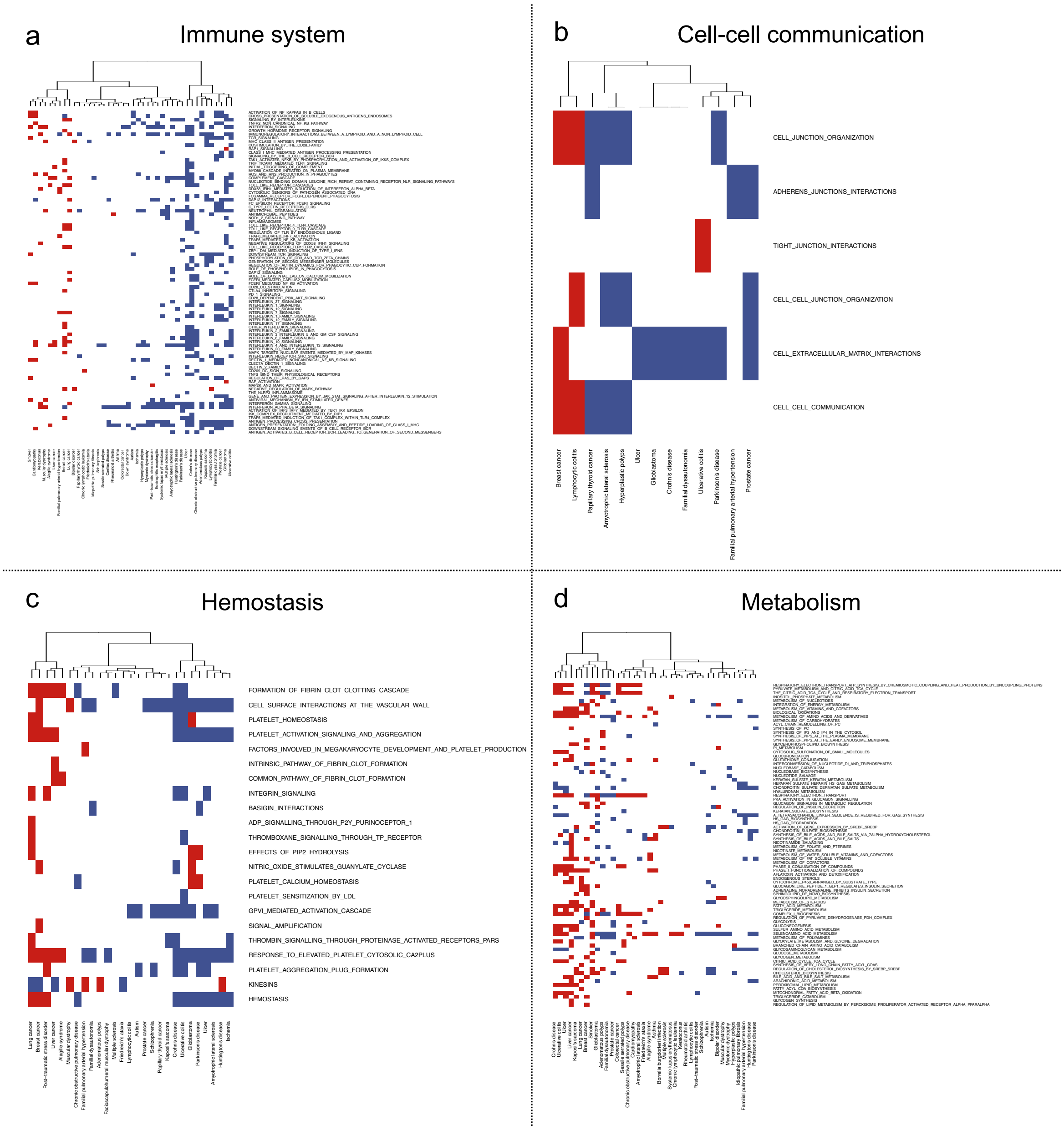


**Fig. S4. Reactome pathways significantly enriched in human diseases: immune system, cell-cell communication, hemostasis, and metabolism.** Reactome pathways significantly over and underexpressed for each disease were identified using GSEA (54) method (FDR < 0.05). Ward2 algorithm was applied to cluster diseases based on the Euclidean distance of the binarized Normalized Effect Size (Methods). Each heatmap shows the significantly altered pathways within the pathway category (rows) across diseases (columns), where over and underexpressed pathways are represented by blue and red colors, respectively. Only diseases with dysregulated pathways are shown. Subpanels depict enriched pathways related to (**a**) Immune system, (**b**) Cell-cell communication, (**c**) Hemostasis, and (**d**) Metabolism.

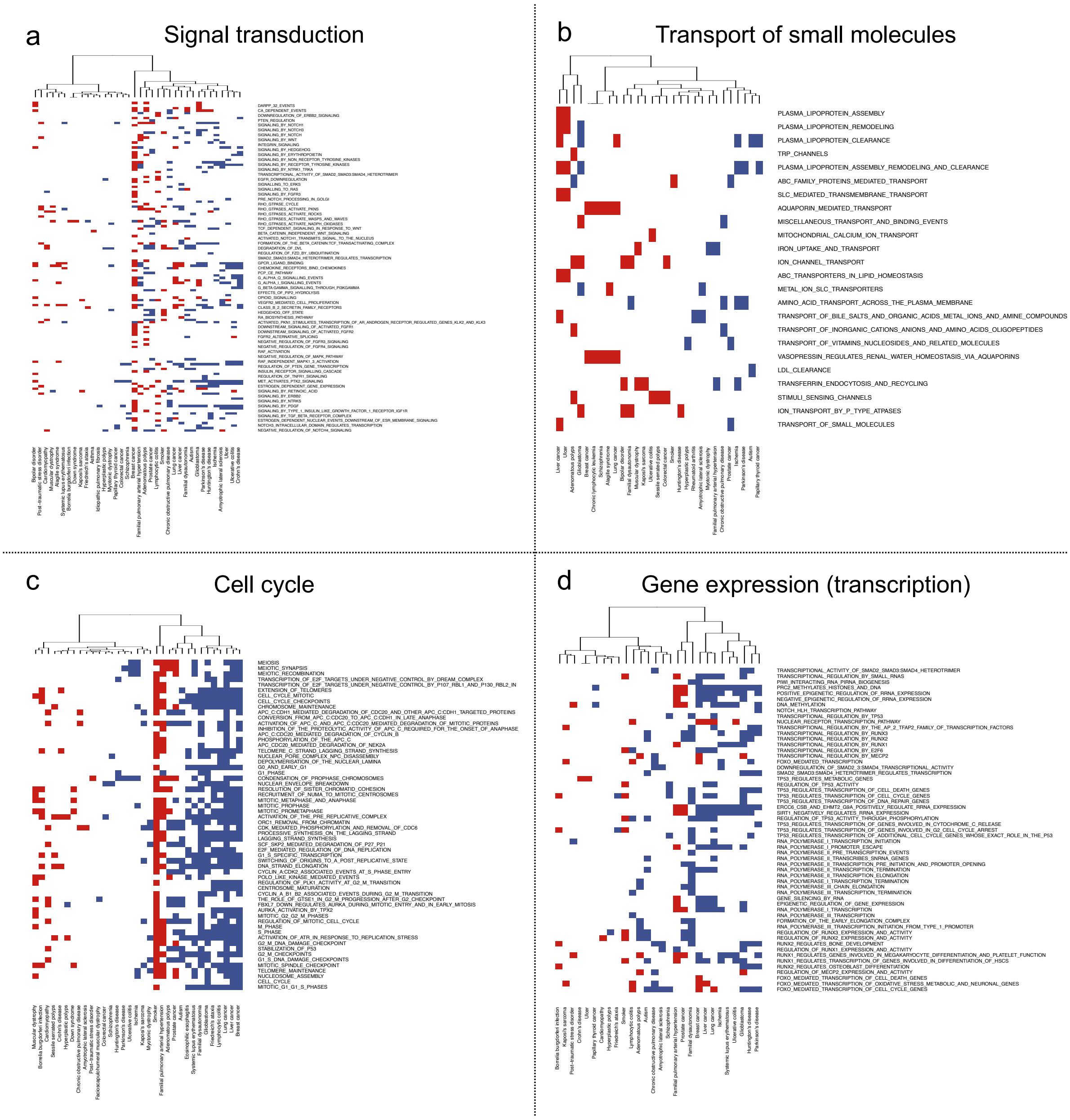


**Fig. S5. Reactome pathways significantly enriched in human diseases: signal transduction, transport of small molecules, cell cycle, and gene expression.** Reactome pathways significantly over and underexpressed for each disease were identified using GSEA (54) method (FDR < 0.05). Ward2 algorithm was applied to cluster diseases based on the Euclidean distance of the binarized Normalized Effect Size (Methods). Each heatmap shows the significantly altered pathways within the pathway category (rows) across diseases (columns), where over and underexpressed pathways are represented by blue and red colors, respectively. Only diseases with dysregulated pathways are shown. Subpanels depict enriched pathways related to (**a**) Signal transduction, (**b**) Transport of small molecules, (**c**) Cell cycle, and (**d**) Gene expression.

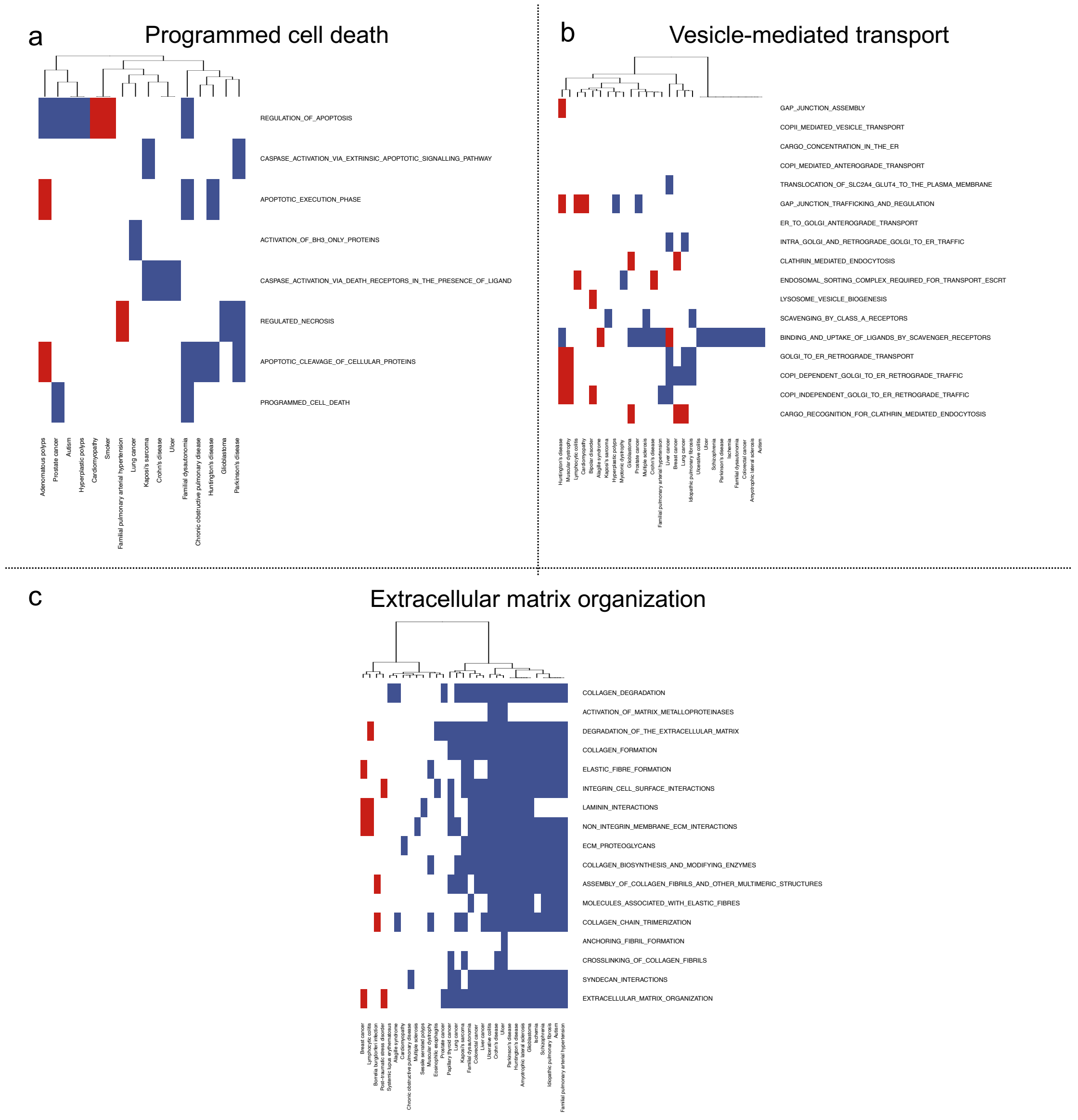


**Fig. S6. Reactome pathways significantly enriched in human diseases: programmed cell death, vesicle-mediated transport, and extracellular matrix organization.** Reactome pathways significantly over and underexpressed for each disease were identified using GSEA (54) method (FDR < 0.05). Ward2 algorithm was applied to cluster diseases based on the Euclidean distance of the binarized Normalized Effect Size (Methods). Each heatmap shows the significantly altered pathways within the pathway category (rows) across diseases (columns), where over and underexpressed pathways are represented by blue and red colors, respectively. Only diseases with dysregulated pathways are shown. Subpanels depict enriched pathways related to (**a**) Programmed cell death, (**b**) Vesicle-mediated transport, and (**c**) Extracellular matrix organization.

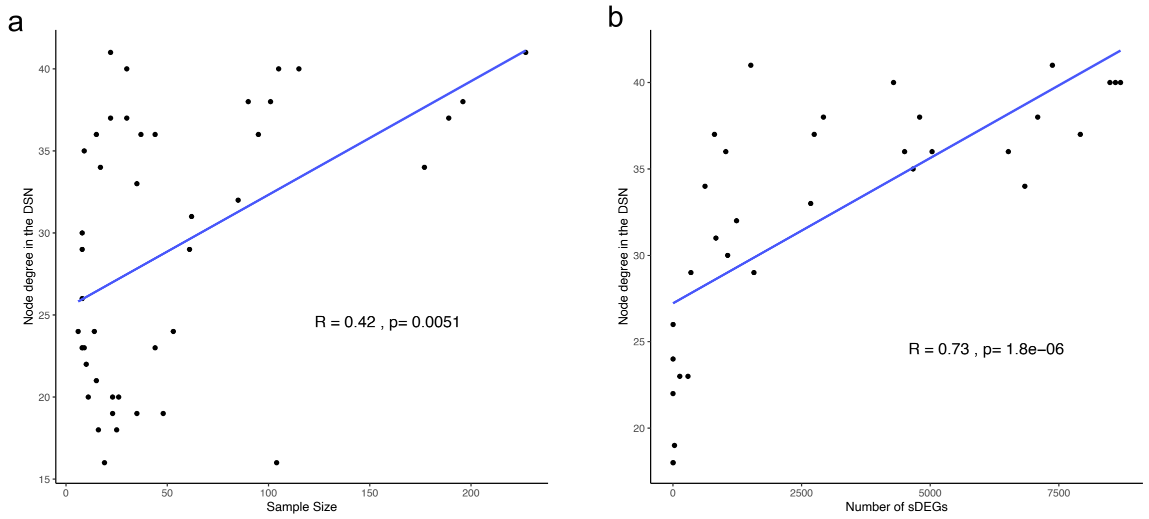


**Fig. S7. Correlation between the diseases' node degree in the Disease Similarity Network (DSN) with sample size and the number of sDEGs.** (**a**) Spearman's correlation between the node degree of the diseases in the DSN and their respective sample size. (**b**) Spearman's correlation between the node degree of the diseases in the DSN and their number of significantly differentially expressed genes (sDEGs).

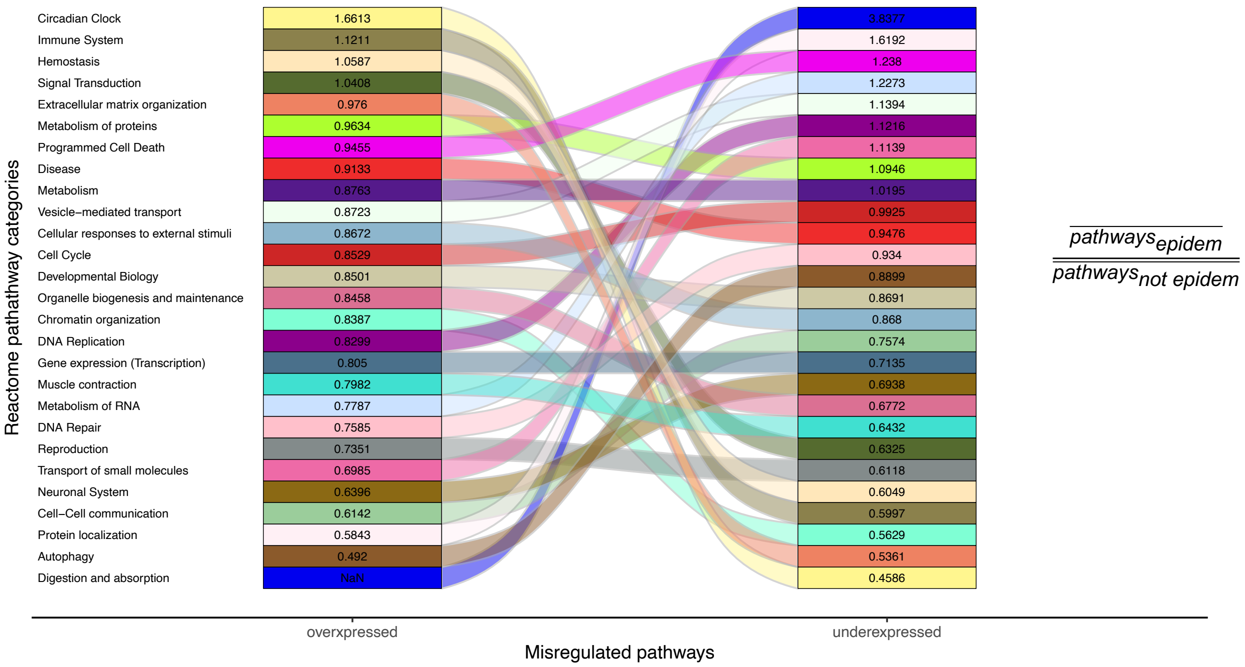


**Fig. S8. Sankey plot illustrating the pathway categories commonly over and underexpressed in epidemiological versus non-epidemiological interactions.** Each color in the plot corresponds to a Reactome pathway category (rows). Pathway categories are sorted based on the ratio between the mean number of shared pathways in epidemiological versus non-epidemiological interactions for over and underexpressed pathways (columns).

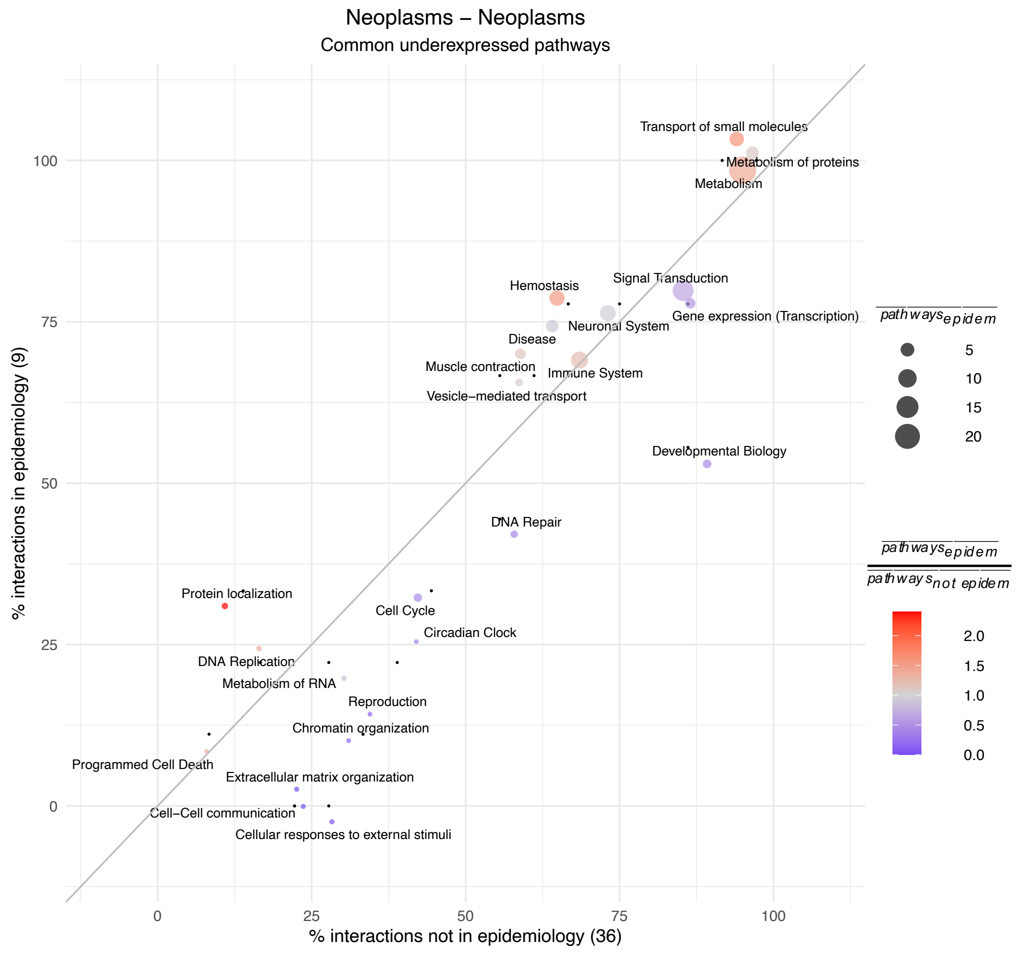


**Fig. S9. Underexpressed pathways behind epidemiological and non-epidemiological interactions between neoplasms.** Percentage of epidemiological (EIs) versus non-epidemiological interactions (NEIs) between neoplasms sharing underexpressed pathways. Each data point represents a Reactome pathway category, with point size indicating the mean number of shared pathways in EIs. Point color reflects the ratio of the mean number of shared underexpressed pathways in EIs versus NEIs (e.g. red indicates that epidemiological interactions share more pathways than non-epidemiological interactions). The number of EIs and NEIs is specified between parentheses in the y and x-axis labels, respectively.

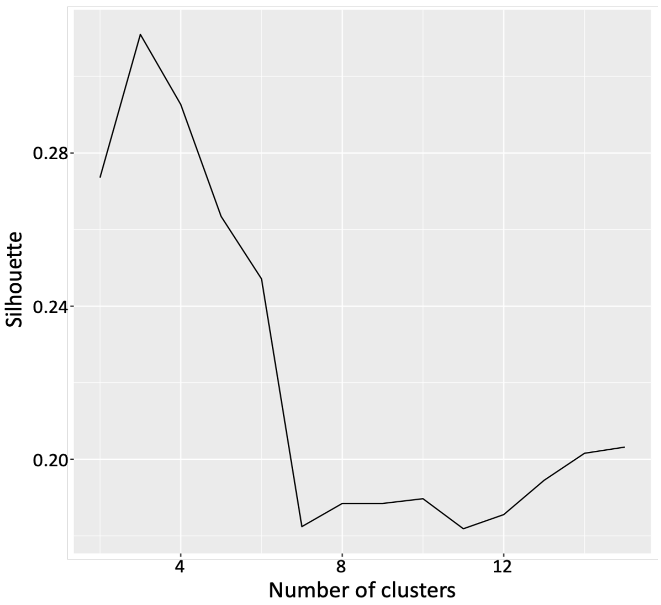


**Fig. S10. Silhouette values in breast cancer patients' clustering.** Silhouette values obtained by applying PAM algorithm to cluster breast cancer patients using a range of cluster numbers from 2 to 15.

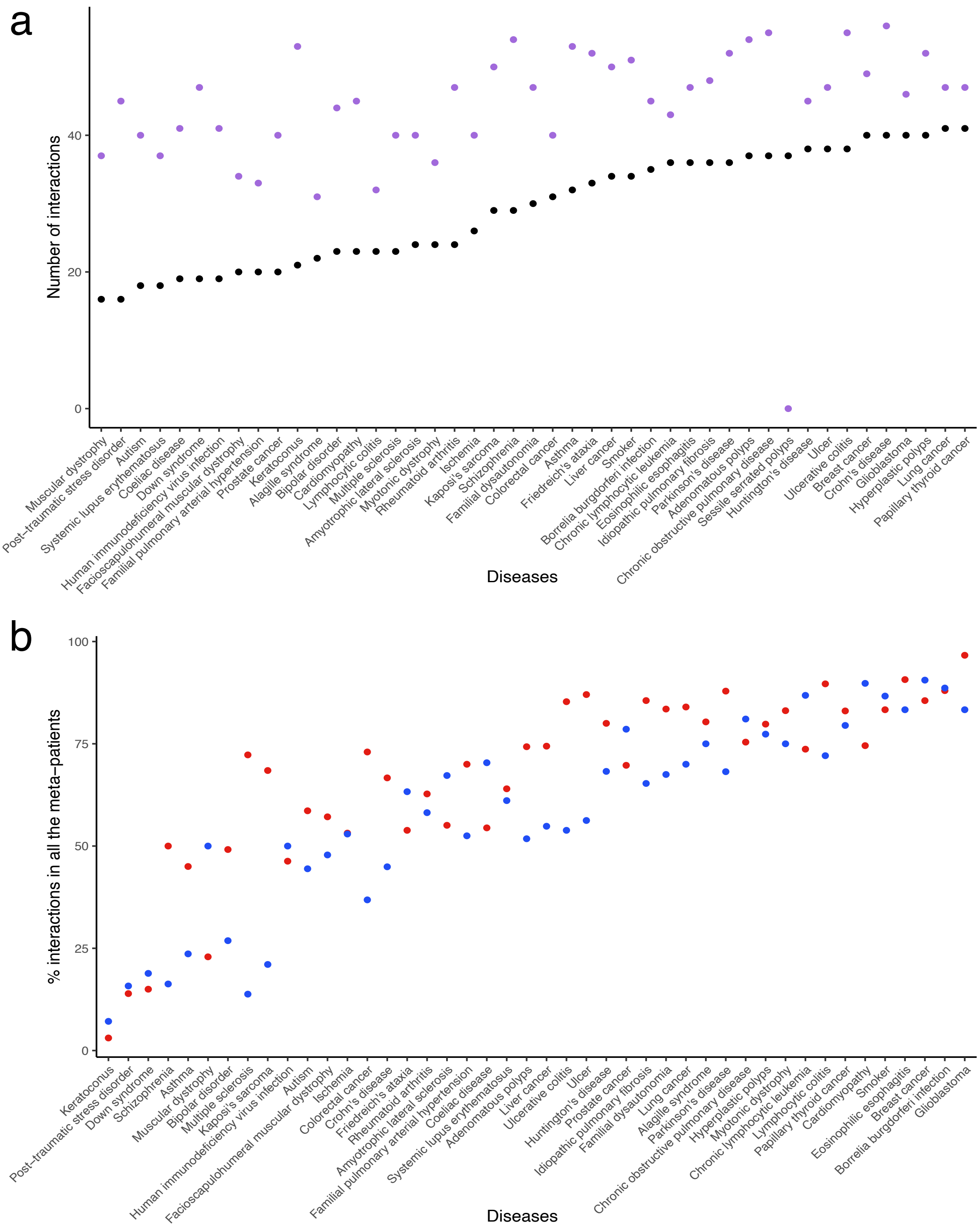


**Fig. S11. Comparison of the total number of interactions at the disease and meta-patient levels and heterogeneity of interactions at the meta-patient level.** (**a**) Total number of interactions for a given disease at both the disease and meta-patient levels. For each disease (x-axis), it is represented the total number of interactions with unique diseases at the disease level (black) and the meta-patient level (purple). Diseases are sorted based on the number of interactions at the disease level. Meta-patients were considered linked to a given disease if they were connected to the disease itself or one of the disease meta-patients. Interactions between meta-patients or diseases from the same disease were omitted for clarity. (**b**) Percentage of interactions observed for all the diseases' meta-patients. For each disease (x-axis), it is represented the percentage of positive (red) and negative (blue) interactions with unique diseases present in all its meta-patients. Diseases are sorted by the mean of their percentage considering the positive and negative interactions. Again, meta-patients were considered linked to a given disease if connected to the disease itself or one of the disease meta-patients, with interactions between meta-patients or diseases from the same disease omitted for clarity.

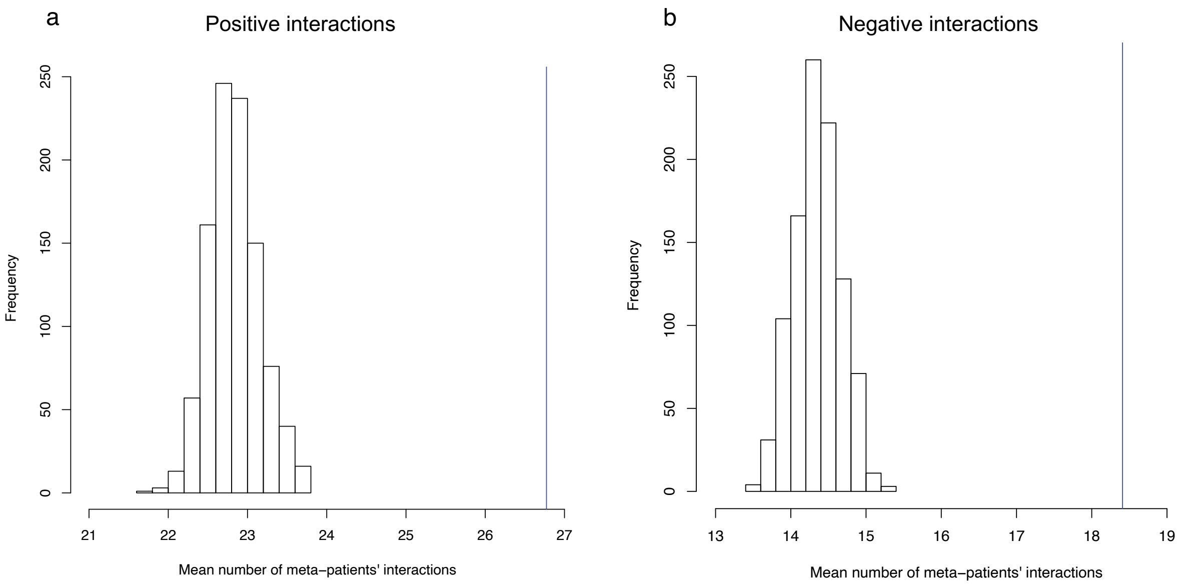


**Fig. S12. Histograms of the mean number of meta-patients' interactions**. Histograms of the mean number of meta-patients' (**a**) positive and (**b**) negative interactions obtained through randomizations (Methods). The blue line represents the number of interactions derived from the Stratified Similarity Network (SSN).

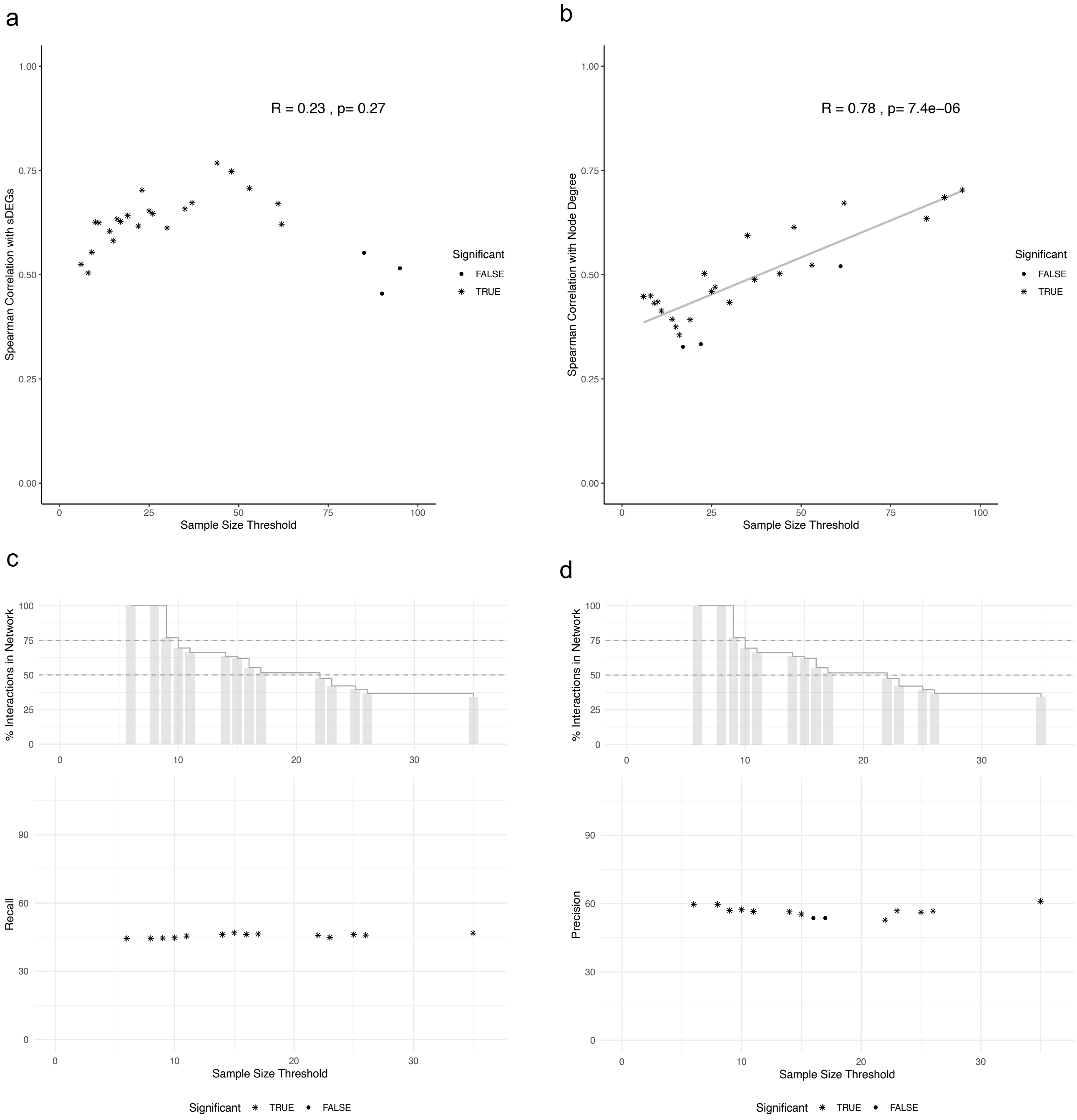


**Fig. S13. Saturation analysis.** *(Top panels)* Scatter plots showing the correlation between sample size and (**a**) the number of significantly differentially expressed genes (sDEGs) or (**b**) node degree in the Disease Similarity Network (DSN) as the sample size threshold increases, progressively removing diseases with lower sample sizes. Statistically significant Spearman's correlations are marked with asterisks. The overall Spearman's correlation coefficient (R) and p-value are reported in each panel. *(Bottom panels)* (**c**) Recall and (**d**) Precision of the DSN compared to epidemiological interactions from Hidalgo et al., (1) evaluated after applying increasing sample size thresholds (Methods). Statistically significant precision and recall values are marked with asterisks. The top bar plots indicate the proportion of interactions retained in the DSN after filtering out diseases below each sample size threshold, with dashed horizontal lines marking 75% and 50% of the network size.

**Table S1. Overlap of the DSN with the epidemiology using the alternative approach.** Table containing the overlap of the Disease Similarity Network (DSN) with the epidemiological network from Hidalgo et al. (1) by grouping the diseases that correspond to the same ICD9 code (Note S2). It shows the number of nodes, number of overlapping interactions (overlap), percentage of overlap, and p-value with respect to the DSN and with respect to the epidemiology for both the positive and the negative interactions (rows).

| Type | Characteristics | DSN |
|---|---|---|
| Positive | Number of nodes in ICD9 | 41 |
| | Overlap | 129 |
| | Perc. overlap from DSN | 42.43 |
| | p-value | 0.0236 |
| | Perc. overlap from epidemiology | 39.21 |
| | p-value | 0.0082 |
| Negative | Number of nodes in ICD9 | 41 |
| | Overlap | 58 |
| | Perc. overlap from DSN | 36.48 |
| | p-value | 0.8891 |
| | Perc. overlap from epidemiology | 17.63 |
| | p-value | 0.9229 |

**Table S2. Classification of breast cancer patients using PAM and Ward2 algorithms.** Table containing the distribution of breast cancer patient among the clusters obtained with PAM and Ward2 algorithms, along with their correspondence with disease subtypes (Methods). Columns contain the obtained clusters. Rows correspond to molecular disease subtypes: triple negative (TN), estrogen receptor positive (ER+) and estrogen receptor negative (ER-). Ward's second cluster is divided into its two branches.

| | Cluster 1 | Cluster 2 | Cluster 3 | |
|---|---|---|---|---|
| Triple negative (TN) | 16 | 2 | | **PAM** |
| Estrogen + (ER+) | | 27 | 3 | |
| Estrogen (ER-) | | 3 | 7 | |
| | Cluster 1 | Cluster 2 | | **Ward2** |
| | | *Branch 1* | *Branch 2* | |
| Triple negative (TN) | 16 | 2 | | |
| Estrogen + (ER+) | | 24 | 5 | |
| Estrogen - (ER-) | | 1 | 8 | |

**Table S3. Percentage of interactions in the DSN and the SSN that are included in their respective network backbones.** This table presents the percentage of interactions in the Disease Similarity Network (DSN) and the Stratified Similarity Network (SSN) included in their respective network backbones. The network backbones were determined using the method from Simas et al. (12) and based on the metric and ultra-metric closures (rows) for the entire network and the subnetworks containing only positive or negative interactions (columns).

| | DSN | | | SSN | | |
|---|---|---|---|---|---|---|
| **Interactions** | **All** | **Positive** | **Negative** | **All** | **Positive** | **Negative** |
| Metric (%) | 96.0 | 95.2 | 97.5 | 45.0 | 19.6 | 92.3 |
| Ultra-metric (%) | 13.4 | 10.6 | 18.3 | 3.7 | 2.9 | 5.2 |

**Table S4. Number of genes that are significantly differentially expressed per disease.** Table containing the number of significantly differentially expressed genes (sDEGs) for each disease (rows) (Methods). It displays the number of total, overexpressed and underexpressed genes (total, over, under) in separate columns (Methods).

| | sDEGs | | |
|---|---|---|---|
| **Disease** | **Total** | **Over** | **Under** |
| Adenomatous polyps | 807 | 515 | 292 |
| Alagille syndrome | 1 | 0 | 1 |
| Amyotrophic lateral sclerosis | 2 | 2 | 0 |
| Asthma | 1236 | 647 | 589 |
| Autism | 5 | 3 | 2 |
| Bipolar disorder | 0 | 0 | 0 |
| Borrelia burgdorferi infection | 4668 | 2296 | 2372 |
| Breast cancer | 8493 | 4196 | 4297 |
| Cardiomyopathy | 133 | 57 | 76 |
| Chronic lymphocytic leukemia | 1027 | 513 | 514 |
| Chronic obstructive pulmonary disease | 7918 | 3903 | 4015 |
| Coeliac disease | 0 | 0 | 0 |
| Colorectal cancer | 835 | 316 | 519 |
| Crohn's disease | 8600 | 3879 | 4721 |
| Downs syndrome | 30 | 29 | 1 |
| Eosinophilic esophagitis | 5036 | 2448 | 2588 |
| Facioscapulohumeral dystrophy | 0 | 0 | 0 |
| Familial dysautonomia | 1063 | 355 | 708 |
| Familial pulmonary arterial hypertension | 0 | 0 | 0 |
| Friedreich's ataxia | 2677 | 1331 | 1346 |
| Glioblastoma | 8699 | 4585 | 4114 |
| HIV | 0 | 0 | 0 |
| Huntington's disease | 2925 | 1603 | 1322 |
| Hyperplastic polyps | 4287 | 2231 | 2056 |
| Idiopathic pulmonary fibrosis | 6519 | 3210 | 3309 |
| Ischemia | 3 | 2 | 1 |
| Kaposi's Sarcoma | 1574 | 728 | 846 |
| Keratoconus | 0 | 0 | 0 |
| Liver cancer | 6839 | 3712 | 3127 |
| Lung cancer | 7375 | 3691 | 3684 |
| Lymphocytic colitis | 0 | 0 | 0 |
| Multiple sclerosis | 292 | 29 | 263 |
| Muscular Dystrophy | 0 | 0 | 0 |
| Myotonic Dystrophy | 0 | 0 | 0 |
| Parkinson's disease | 4502 | 2187 | 2315 |
| Posttraumatic stress disorder | 0 | 0 | 0 |
| Prostate cancer | 0 | 0 | 0 |
| Rheumatoid arthritis | 0 | 0 | 0 |
| Systemic lupus erythematosus | 1 | 0 | 1 |
| Schizophrenia | 349 | 175 | 174 |
| Sessile serrated polyposis | 2746 | 1578 | 1168 |
| Smoker | 623 | 509 | 114 |
| Thyroid cancer papillary | 1513 | 622 | 891 |
| Ulcer | 4794 | 2375 | 2419 |
| Ulcerative colitis | 7089 | 3341 | 3748 |

**Table S5. Disease Similarity Network (DSN) properties.** Table containing the general characteristics of the generated disease-disease networks (columns). Properties are provided for the DSN at the disease and ICD9 level, and for the epidemiological network from Hidalgo et al. over the common set of diseases (1), considering all interactions, only positive, and only negative interactions (rows) (Methods). It shows the number of nodes, the number of possible (possible) and detected (significant) interactions, percentage of positive and negative interactions, number of connected components (CC), size of the largest connected component (size CC) and mean degree of the networks (Methods).

| Type | Characteristics | DSN | DSN (ICD9) | Epidemiology |
|---|---|---|---|---|
| All | Number of nodes | 45 | 41 | 41 |
| | Possible | 990 | 820 | 780 |
| | Significant | 658 | 545 | 331 |
| | CC | 1 | 1 | 1 |
| | Size CC | 45 | 41 | 40 |
| | Mean degree | 29.24 | 26.58 | 16.55 |
| Positive | Detected | 417 | 347 | 331 |
| | Percentage | 63.37 | 63.67 | 100 |
| | CC | 1 | 1 | 1 |
| | Biggest CC | 45 | 41 | 40 |
| | Mean degree | 18.53 | 16.93 | 16.55 |
| Negative | Detected | 241 | 198 | |
| | Percentage | 36.63 | 36.33 | |
| | CC | 1 | 1 | |
| | Size CC | 45 | 41 | |
| | Mean degree | 10.71 | 9.66 | |

**Table S6. Overlap of the ICD9 DSN with the epidemiology.** Table containing the overlap of the ICD9 Disease Similarity Network (DSN) with the epidemiological network from Hidalgo et al. (1) (Methods). It shows the number of nodes, number of nodes also in the epidemiology, number of edges entailing the common nodes in the DSN and the epidemiology, number of overlapping interactions (overlap), percentage of overlap and p-value with respect to the epidemiology (recall) and the DSN (precision) for positive and negative interactions (rows).

| Type | Characteristics | DSN |
|---|---|---|
| Positive | Number of nodes in ICD9 | 41 |
| | Nodes in epidem. | 40 |
| | Edges in DSN | 347 |
| | Edges in epidem. | 329 |
| | Overlap | 152 |
| | Perc. overlap from epidemiology | 46.20 |
| | p-value | 0.0018 |
| | Perc. overlap from DSN | 43.80 |
| | p-value | 0.004 |
| Negative | Number of nodes in ICD9 | 41 |
| | Nodes in epidem. | 40 |
| | Edges in DSN | 198 |
| | Edges in epidem. | 329 |
| | Overlap | 73 |
| | Perc. overlap from epidemiology | 22.18 |
| | p-value | 0.867 |
| | Perc. overlap from DSN | 36.87 |
| | p-value | 0.86 |

**Table S7. Overlap of the ICD9 DSN obtained using Spearman's correlation and cosine similarity with the epidemiology.** Table containing the overlap of the ICD9 Disease Similarity Network (DSN) obtained using Spearman's correlation and cosine similarity with the epidemiological network from Hidalgo et al. (1) (Methods). It shows the number of overlapping interactions (overlap), recall (percentage of epidemiological interactions captured by the molecular networks) and precision (percentage of molecular interactions contained in the epidemiological networks), and their respective p-values. Asterisks indicate significant precisions and recalls.

| | Network | Overlap | Recall (%) | p-value | Precision (%) | p-value |
|---|---|---|---|---|---|---|
| DSN | Spearman's correlation | 152 | 46.2* | 0.0018 | 43.8* | 0.004 |
| | Cosine similarity | 162 | 49.24* | 0.0034 | 44.38* | 0.0021 |

**Table S8. Overlap of the ICD9 Disease Similarity Network (DSN) and the Stratified Similarity Network (SSN) with the epidemiology.** Table containing the overlap of the ICD9 Disease Similarity Network (DSN) with the epidemiological network from Hidalgo et al. based on Relative Risks (RR) and phi-correlation (1) (Methods). It shows the number of overlapping interactions (overlap), recall (percentage of epidemiological interactions captured by the molecular networks) and precision (percentage of molecular interactions contained in the epidemiological networks), and their respective p-values. Asterisks indicate significant precisions and recalls.

| | Network | Overlap | Recall (%) | p-value | Precision (%) | p-value |
|---|---|---|---|---|---|---|
| DSN | RR | 152 | 46.2* | 0.0018 | 43.8* | 0.004 |
| | Phi-correlation | 207 | 44.4* | 0.0016 | 59.7* | 0.0018 |
| SSN | RR | 211 | 64.13* | 0.0187 | 43.06* | 0.0253 |
| | Phi-correlation | 294 | 63.1* | 0.0025 | 60* | 0.0034 |

**Table S9. Topological properties of the ICD9 Disease Similarity Network (DSN) and the comparable epidemiological subnetwork.** Table containing general topological properties of the ICD9 DSN entailing positive interactions and the epidemiological network by Hidalgo et al. (1) over the common set of diseases (columns). It includes information on the number (CC) and size (size CC) of the connected components, number of interactions (N interactions), mean degree, density, mean distance (mean length of the shortest paths), diameter (longest shortest path), mean transitivity, mean closeness, mean betweenness, mean degeneracy and disease category assortativity (rows). The assortativity was computed by labelling nodes with their corresponding disease category. The edge weight was not considered for these calculations. When applicable, a paired t-test was used to assess the significance of the differences between the mean topological values of the two networks (considering as the null hypothesis that the means are equal).

| | | **DSN (common ICD9)** | **Epidemiology (common ICD9)** | **t-test p-value** |
|---|---|---|---|---|
| Positive | CC | 1 | 1 | - |
| | Size CC | 40 | 40 | - |
| | N interactions | 327 | 329 | - |
| | Mean degree | 16.35 | 16.45 | 0.953 |
| | Density | 0.419 | 0.422 | - |
| | Mean distance | 1.588 | 1.665 | - |
| | Diameter | 3 | 4 | - |
| | Mean transitivity | 0.556 | 0.664 | 0.001 |
| | Mean closeness | 0.0163 | 0.016 | 0.423 |
| | Mean betweenness | 11.475 | 12.975 | 0.605 |
| | Mean degeneracy | 10.825 | 10.95 | 0.845 |
| | Dis. Categ. Assortativity | 0.035 | -0.036 | - |

**Table S10. Overlap of other disease-disease networks based on molecular information with the epidemiology.** This table presents the number of nodes, edges and overlaps of positive interactions in disease-disease networks based on microbiome (55), miRNA (56), PPI (4), cellular components -PPIs and genes- (6), and subcellular localization (5) (columns). It shows the number of nodes and edges of the original networks (positive interactions). Then, it provides the number of nodes and edges of the molecular networks transformed into ICD9 codes (all of them in the epidemiology), the number of edges of the epidemiological network involving these nodes, the number of overlapping interactions (overlap), percentage, and p-value of the overlaps with the epidemiological network from Hidalgo et al. (1) (ICD9 positive interactions). Additionally, it shows the above information after selecting only the ICD9 codes present in the Disease Similarity Network (DSN) (ICD9 positive interactions in the DSN) (Methods).

| Type | Characteristics | Microbiome | miRNA | PPI | Cellular components (PPIs) | Cellular components (genes) | Subcellular localization |
|---|---|---|---|---|---|---|---|
| Positive interactions | Number of nodes | 33 | 63 | 289 | 379 | 379 | 397 |
| | Number of edges | 112 | 414 | 1383 | 1873 | 658 | 7584 |
| ICD9 positive interactions | Number of nodes | 23 | 46 | 136 | 202 | 193 | 211 |
| | Nodes in epidem. | 23 | 46 | 136 | 202 | 193 | 211 |
| | Number of edges | 87 | 324 | 536 | 1509 | 503 | 4934 |
| | Edges in epidem. | 169 | 358 | 4593 | 9921 | 8983 | 10283 |
| | Overlap | 61 | 129 | 400 | 658 | 293 | 2431 |
| | Perc. overlap from molecular network | 70.11 | 39.81 | 74.63 | 43.61 | 58.25 | 49.27 |
| | p-value | 0.225 | 0.265 | 0 | 0.0059 | 0 | 0.3234 |
| | Perc. overlap from epidemiology | 36.09 | 36.03 | 8.71 | 6.63 | 3.26 | 23.64 |
| | p-value | 0.067 | 0.412 | 0 | 0.0009 | 0 | 0.2692 |
| ICD9 positive interactions in the DSN | Nodes in epidem. | 6 | 11 | 19 | 26 | 25 | 26 |
| | Number of edges | 8 | 40 | 20 | 104 | 37 | 108 |
| | Edges in epidem. | 11 | 16 | 81 | 98 | 98 | 107 |
| | Overlap | 7 | 9 | 15 | 26 | 11 | 39 |
| | Perc. overlap from molecular network | 87.5 | 22.5 | 75 | 25 | 29.73 | 36.11 |
| | p-value | 1 | 0.961 | 0.0031 | 0.2649 | 0.0566 | 0.542 |
| | Perc. overlap from epidemiology | 63.64 | 56.25 | 18.52 | 26.53 | 11.22 | 36.45 |
| | p-value | 1 | 0.96 | 0.0045 | 0.1966 | 0.0095 | 0.639 |

**Table S11. Comparison of the Disease Similarity Network (DSN, RNA-seq) with the Disease Molecular Similarity Network (microarrays) and the disease network derived from PPI.** Table containing the number of nodes and edges, along with the number of common nodes and number of edges within common nodes for the DSN and the microarrays network by Sánchez-Valle et al. (22) or the network based on PPI by Menche et al. (4). For the comparison, all the network nodes were transformed into ICD9 codes.

| | **Comparison with microarrays** | | **Comparison with PPI** | |
|---|---|---|---|---|
| | DSN (ICD9) | Microarrays (ICD9) | DSN (ICD9) | PPI (ICD9) |
| Number of nodes | 41 | 92 | 41 | 289 |
| Number of edges | 545 | 2155 | 545 | 1383 |
| Common nodes | 27 | 27 | 19 | 19 |
| Number of edges within common nodes | 251 | 134 | 73 | 20 |

**Table S12. Overlap between the Disease Similarity Network (DSN, RNAseq) with the Disease Molecular Similarity Network (microarrays) and the disease network derived from PPI.** Table containing the overlap between the ICD9 DSN with the microarray network from Sánchez-Valle et al. (22) and the PPI-based network derived from Menche et al. (4) (columns). The overlapping is provided for all the interactions, the positive and negative ones when applicable (rows) (Methods). The table includes the overlap (number of common interactions), the percentage (percentage of interactions from the molecular networks captured by the DSN) and significance of the overlap (Methods).

| Type | Characteristics | DSN (ICD9) vs microarrays | DSN (ICD9) vs PPI |
|---|---|---|---|
| All | Overlap | 63 | 6 |
| | Percentage overlap | 47.015% | 30% |
| | p-value | 0.0272 | 0.942 |
| Positive | Overlap | 45 | 6 |
| | Percentage overlap | 65.217% | 30% |
| | p-value | 0.002 | 0.942 |
| Negative | Overlap | 18 | - |
| | Percentage overlap | 27.692% | - |
| | p-value | 0.6243 | - |

**Table S13. Top immune system pathways from Reactome involved in epidemiological interactions.** This table displays the top 20 immune system pathways from Reactome (23) that are commonly and significantly overexpressed in epidemiological interactions (EIs) of the Disease Similarity Network (DSN). It includes the name of the pathway and the number of EIs that share its significant overexpression.

| Top Immune system Reactome pathways | Number of EIs |
|---|---|
| Interferon alpha beta signaling | 51 |
| Antigen presentation: folding assembly and peptide loading of class I MHC | 51 |
| Interleukin 4 and interleukin 13 signaling | 41 |
| Antigen processing cross presentation | 34 |
| Complement cascade | 33 |
| Interferon gamma signaling | 33 |
| Interferon signaling | 27 |
| Immunoregulatory interactions between a lymphoid and a non-lymphoid cell | 25 |
| Neutrophil degranulation | 25 |
| DAP12 interactions | 16 |
| Interleukin 2 family signaling | 9 |
| Interleukin 3 interleukin 5 and GM-CSF signaling | 9 |
| Interleukin 6 family signaling | 9 |
| Interleukin receptor SHC signaling | 9 |
| Signaling by interleukins | 8 |
| Costimulation by the CD28 family | 8 |
| ROS and RNS production in phagocytes | 8 |
| Interleukin 1 signaling | 8 |
| Interleukin 1 family signaling | 8 |
| Interleukin 10 signaling | 8 |

**Table S14. Topological properties of the Disease Similarity Network (DSN) and the Stratified Similarity Network (SSN).** Table containing general and topological properties of the DSN and the SSN (columns). Properties are provided for all the interactions, the positive and the negative ones, when applicable (rows). It shows the number (CC) and size (size CC) of the connected components, mean degree, density, mean transitivity, diameter (longest shortest path) and mean distance (mean length of the shortest paths). Edge weight was not considered in these calculations.

| | | **DSN** | **SSN** |
|---|---|---|---|
| All | CC | 1 | 1 |
| | Size CC | 45 | 161 |
| | Mean degree | 29.244 | 112.012 |
| | Density | 0.665 | 0.700 |
| | Mean transitivity | 0.705 | 0.746 |
| | Diameter | 2 | 2 |
| | Mean distance | 1.335 | 1.300 |
| Positive | CC | 1 | 1 |
| | Size CC | 45 | 161 |
| | Mean degree | 18.533 | 72.907 |
| | Density | 0.421 | 0.456 |
| | Mean transitivity | 0.564 | 0.611 |
| | Diameter | 3 | 3 |
| | Mean distance | 1.587 | 1.545 |
| Negative | CC | 1 | 1 |
| | Size CC | 45 | 161 |
| | Mean degree | 10.711 | 39.106 |
| | Density | 0.243 | 0.244 |
| | Mean transitivity | 0.156 | 0.151 |
| | Diameter | 3 | 3 |
| | Mean distance | 1.825 | 1.772 |

**Table S15. Overlap of the interactions at the meta-patient level with the epidemiology.** Table containing the overlap of the interactions between meta-patients and diseases with the epidemiological network from Hidalgo et al. (1) (Methods). It shows the number of overlapping interactions (overlap), percentage of overlap, and p-value with respect to the network (precision) and with respect to the epidemiology (recall) for both positive and negative interactions (rows).

| Type | Characteristics | Meta-patient level |
|---|---|---|
| Positive | Overlap | 211 |
| | Perc. overlap from DSN | 43.06 |
| | p-value | 0.0253 |
| | Perc. overlap from epidemiology | 64.13 |
| | p-value | 0.0187 |
| Negative | Overlap | 135 |
| | Perc. overlap from DSN | 41.28 |
| | p-value | 0.8082 |
| | Perc. overlap from epidemiology | 40.79 |
| | p-value | 0.8035 |

**Table S16. Literature validation of false positives in the Disease Similarity Network (DSN).** Table showing the literature validation of the top 100 and bottom 100 interactions initially labeled as false positives in the DSN, after the comparison with the epidemiological network from Hidalgo et al. (1) based on relative risks (RR) (Methods). Interactions were ranked by correlation strength, and only unique ICD9 disease pairs are included. These account for 71.4% of the total number of false positives in the DSN. The table lists the disease pairs involved in each interaction. The column (In epidemiology) indicates whether the interaction was supported by epidemiological evidence, including the phi-correlation network from Hidalgo et al. or recent large-scale studies. The table also reports the source of supporting evidence (References) and the corresponding citation numbers (Ref. IDs). A thick horizontal line separates the top and bottom ranked interactions.

| Disease 1 | Disease 2 | In epidemiology | References | Ref. IDs |
|---|---|---|---|---|
| Amyotrophic lateral sclerosis | Alagille syndrome | True | phi-correlation | (1) |
| Amyotrophic lateral sclerosis | Keratoconus | - | | |
| Prostate cancer | Systemic lupus erythematosus | True | (Ou et al., 2023) | (57) |
| Lymphocytic colitis | Prostate cancer | - | | |
| Muscular dystrophy | Prostate cancer | True | (Maya-González et al., 2024) | (58) |
| Post-traumatic stress disorder | Prostate cancer | True | (Anastasiou et al., 2011; Clouston et al., 2019) | (59, 60) |
| Bipolar disorder | Prostate cancer | True | (Chen et al., 2015; Chrobak et al., 2023) | (61, 62) |
| Keratoconus | Prostate cancer | - | | |
| Amyotrophic lateral sclerosis | Prostate cancer | - | | |
| Ischemia | Autism | True | phi-correlation | (1) |
| Down syndrome | Bipolar disorder | True | phi-correlation | (1) |
| Multiple sclerosis | Huntington's disease | True | phi-correlation | (1) |
| Autism | Systemic lupus erythematosus | - | | |
| Down syndrome | Systemic lupus erythematosus | True | phi-correlation | (1) |
| Huntington's disease | Glioblastoma | True | phi-correlation | (1) |
| Parkinson's disease | Glioblastoma | True | (Cedergren Weber et al., 2023; Leong et al., 2021; Park, 2019; Ye et al., 2016) | (46–49) |
| Amyotrophic lateral sclerosis | Rheumatoid arthritis | - | | |
| Lung cancer | Breast cancer | True | (Curtis RE et al., 2006) | (7) |
| Glioblastoma | Bipolar disorder | True | phi-correlation | (1) |
| Coeliac disease | Ischemia | True | phi-correlation | (1) |
| Amyotrophic lateral sclerosis | Autism | True | (Chuquilin et al., 2017; O'Brien et al., 2017) | |
| Multiple sclerosis | Parkinson's disease | - | | |
| Cardiomyopathy | Rheumatoid arthritis | True | phi-correlation | (1) |
| Adenomatous polyps | Prostate cancer | True | (Ko et al., 2016) | (63) |
| Glioblastoma | Multiple sclerosis | - | - | - |
| Chronic obstructive pulmonary disease | Keratoconus | - | | |
| Smoker | Multiple sclerosis | True | (Nishanth et al., 2020; Wingerchuk, 2012) | (64, 65) |
| Parkinson's disease | Asthma | True | (Nam et al., 2024; Cheng et al., 2015) | (38, 39) |
| Breast cancer | Lymphocytic colitis | - | | |
| Amyotrophic lateral sclerosis | Crohn's disease | - | | |

| | | | | |
|---|---|---|---|---|
| Alagille syndrome | Asthma | True | phi-correlation | (1) |
| Glioblastoma | Schizophrenia | - | | |
| Papillary thyroid cancer | Asthma | - | | |
| Crohn's disease | Kaposi's sarcoma | True | phi-correlation | (1) |
| Ulcer | Kaposi's sarcoma | True | phi-correlation | (1) |
| Adenomatous polyps | Autism | True | phi-correlation | (1) |
| Multiple sclerosis | Schizophrenia | True | (Andreassen et al., 2015; Misiak et al., 2023) | (66, 67) |
| Papillary thyroid cancer | Parkinson's disease | True | (Ejma et al., 2020) | (68) |
| Smoker | Parkinson's disease | - | | |
| Coeliac disease | Down syndrome | True | (Pavlovic et al., 2017; Satgé and Seidel, 2018) | (69, 70) |
| Down syndrome | Smoker | - | | |
| Amyotrophic lateral sclerosis | Ulcerative colitis | True | phi-correlation | (1) |
| Papillary thyroid cancer | Smoker | - | | |
| Multiple sclerosis | Down syndrome | - | | |
| Hyperplastic polyps | Parkinson's disease | - | | |
| Myotonic dystrophy | Down syndrome | True | phi-correlation | (1) |
| Breast cancer | Adenomatous polyps | True | (Abu-Sbeih et al., 2019) | (71) |
| Prostate cancer | Familial dysautonomia | - | | |
| Papillary thyroid cancer | Sessile serrated polyps | - | | |
| Prostate cancer | Idiopathic pulmonary fibrosis | True | (Lee et al., 2021) | (72) |
| Crohn's disease | Colorectal cancer | True | phi-correlation | (1) |
| Breast cancer | Alagille syndrome | - | | |
| Glioblastoma | Autism | - | | |
| Amyotrophic lateral sclerosis | Schizophrenia | True | (Turner et al., 2016; Zucchi et al., 2019) | (73, 74) |
| Breast cancer | Asthma | True | (Nechuta et al., 2013; Santos-Mejías et al., 2024) | (75, 76) |
| Amyotrophic lateral sclerosis | Cardiomyopathy | True | (Choi et al., 2017; Gdynia et al., 2006; Namazi et al., 2014; Rosenbohm et al., 2017; Xu et al., 2022) | (77–81) |
| Cardiomyopathy | Crohn's disease | - | | |
| Papillary thyroid cancer | Ulcer | True | (Lee et al., 2022) | (82) |
| Schizophrenia | Ischemia | - | | |
| Hyperplastic polyps | Down syndrome | - | | |
| Amyotrophic lateral sclerosis | Familial dysautonomia | True | phi-correlation | (1) |
| Alagille syndrome | Human immunodeficiency virus infection | - | | |
| Kaposi's sarcoma | Papillary thyroid cancer | True | phi-correlation | (1) |
| Borrelia burgdorferi infection | Familial pulmonary arterial hypertension | - | | |
| Breast cancer | Post-traumatic stress disorder | True | (Brown et al., 2020) | (83) |
| Liver cancer | Breast cancer | - | | |
| Familial dysautonomia | Idiopathic pulmonary fibrosis | True | phi-correlation | (1) |
| Smoker | Systemic lupus erythematosus | True | (Chua et al., 2020) | (84) |
| Smoker | Keratoconus | - | | |
| Ulcerative colitis | Huntington's disease | True | phi-correlation | (1) |
| Ulcerative colitis | Keratoconus | True | phi-correlation | (1) |
| Breast cancer | Papillary thyroid cancer | True | (Curtis RE et al., 2006; Trinh et al., 2021) | (7, 85) |
| Chronic lymphocytic leukemia | Smoker | True | (Brown et al., 1992; Richardson et al., 2008) | (86, 87) |
| Chronic lymphocytic leukemia | Post-traumatic stress disorder | True | phi-correlation | (1) |
| Breast cancer | Facioscapulohumeral muscular dystrophy | - | | |
| Kaposi's sarcoma | Glioblastoma | True | phi-correlation | (1) |
| Papillary thyroid cancer | Lung cancer | True | phi-correlation | (1) |
| Autism | Multiple sclerosis | True | phi-correlation | (1) |
| Papillary thyroid cancer | Liver cancer | True | phi-correlation | (1) |
| Familial dysautonomia | Ischemia | True | phi-correlation | (1) |
| Colorectal cancer | Prostate cancer | True | (Curtis RE et al., 2006) | (7) |

| | | | | |
|---|---|---|---|---|
| Papillary thyroid cancer | Myotonic dystrophy | True | phi-correlation | (1) |
| Lung cancer | Schizophrenia | True | (Arffman et al., 2019) | (88) |
| Kaposi's sarcoma | Breast cancer | True | (Curtis RE et al., 2006) | (7) |
| Ulcer | Smoker | True | phi-correlation | (1) |
| Lung cancer | Prostate cancer | - | | |
| Huntington's disease | Keratoconus | - | | |
| Human immunodeficiency virus infection | Colorectal cancer | - | | |
| Kaposi's sarcoma | Liver cancer | True | phi-correlation | (1) |
| Colorectal cancer | Smoker | True | (Amitay et al., 2020; Botteri et al., 2008) | (89, 90) |
| Crohn's disease | Keratoconus | True | (Jin et al., 2024; Tréchot et al., 2015) | (91, 92) |
| Multiple sclerosis | Ischemia | True | (LeVine, 2016) | (93) |
| Schizophrenia | Facioscapulohumeral muscular dystrophy | True | phi-correlation | (1) |
| Coeliac disease | Parkinson's disease | True | phi-correlation | (1) |
| Borrelia burgdorferi infection | Keratoconus | True | phi-correlation | (1) |
| Alagille syndrome | Parkinson's disease | - | | |
| Huntington's disease | Ischemia | - | | |
| Lung cancer | Lymphocytic colitis | True | (Bergman et al., 2020) | (94) |
| Multiple sclerosis | Sessile serrated polyps | - | | |
| Lung cancer | Autism | - | | |
| Papillary thyroid cancer | Lymphocytic colitis | - | | |
| Glioblastoma | Colorectal cancer | - | | |
| Glioblastoma | Eosinophilic esophagitis | - | | |
| Papillary thyroid cancer | Idiopathic pulmonary fibrosis | True | (Lee et al., 2021) | (72) |
| Parkinson's disease | Systemic lupus erythematosus | True | (Kim et al., 2023; Mv et al., 2023) | (95, 96) |
| Crohn's disease | Schizophrenia | True | (Benros et al., 2014; Qian et al., 2022) | (97, 98) |
| Down syndrome | Chronic lymphocytic leukemia | True | phi-correlation | (1) |
| Papillary thyroid cancer | Keratoconus | - | | |
| Eosinophilic esophagitis | Keratoconus | - | | |
| Idiopathic pulmonary fibrosis | Friedreich's ataxia | True | phi-correlation | (1) |
| Lung cancer | Keratoconus | - | | |
| Papillary thyroid cancer | Friedreich's ataxia | - | | |
| Asthma | Human immunodeficiency virus infection | True | phi-correlation | (1) |
| Familial dysautonomia | Ulcerative colitis | - | | |
| Crohn's disease | Down syndrome | - | | |
| Eosinophilic esophagitis | Down syndrome | True | phi-correlation | (1) |
| Breast cancer | Down syndrome | - | | |
| Liver cancer | Friedreich's ataxia | True | phi-correlation | (1) |
| Idiopathic pulmonary fibrosis | Breast cancer | - | | |
| Kaposi's sarcoma | Bipolar disorder | True | phi-correlation | (1) |
| Huntington's disease | Kaposi's sarcoma | True | phi-correlation | (1) |
| Systemic lupus erythematosus | Breast cancer | - | | |
| Glioblastoma | Lymphocytic colitis | - | | |
| Borrelia burgdorferi infection | Chronic obstructive pulmonary disease | - | | |
| Ulcer | Human immunodeficiency virus infection | True | phi-correlation | (1) |
| Borrelia burgdorferi infection | Ischemia | True | (Garkowski et al., 2017; Moreno Legast et al., 2018) | (99, 100) |
| Glioblastoma | Coeliac disease | - | | |
| Systemic lupus erythematosus | Schizophrenia | True | (Tiosano et al., 2017) | (101) |
| Familial dysautonomia | Liver cancer | - | | |
| Idiopathic pulmonary fibrosis | Keratoconus | - | | |
| Ulcerative colitis | Borrelia burgdorferi infection | True | phi-correlation | (1) |

| | | | | |
|---|---|---|---|---|
| Amyotrophic lateral sclerosis | Colorectal cancer | - | | |
| Colorectal cancer | Chronic lymphocytic leukemia | True | (Dennis and Alberts, 2007; van der Straten et al., 2023) | (102, 103) |
| Down syndrome | Friedreich's ataxia | True | phi-correlation | (1) |
| Chronic lymphocytic leukemia | Human immunodeficiency virus infection | True | phi-correlation | (1) |
| Eosinophilic esophagitis | Breast cancer | - | | |
| Autism | Breast cancer | True | (Kao et al., 2010) | (104) |
| Multiple sclerosis | Kaposi's sarcoma | True | phi-correlation | (1) |
| Coeliac disease | Schizophrenia | True | phi-correlation | (1) |
| Familial pulmonary arterial hypertension | Friedreich's ataxia | True | (Culley et al., 2023, 2021; Jensen and Bundgaard, 2012) | (105–107) |
| Prostate cancer | Asthma | True | (Su et al., 2015) | (108) |
| Alagille syndrome | Cardiomyopathy | True | phi-correlation | (1) |
| Colorectal cancer | Breast cancer | True | (Curtis RE et al., 2006) | (7) |
| Familial pulmonary arterial hypertension | Parkinson's disease | - | | |
| Papillary thyroid cancer | Eosinophilic esophagitis | True | phi-correlation | (1) |
| Post-traumatic stress disorder | Borrelia burgdorferi infection | True | phi-correlation | (1) |
| Papillary thyroid cancer | Crohn's disease | True | (Wadhwa et al., 2016) | (109) |
| Papillary thyroid cancer | Colorectal cancer | True | (Sandeep et al., 2006) | (110) |
| Coeliac disease | Lung cancer | - | | |
| Kaposi's sarcoma | Schizophrenia | True | phi-correlation | (1) |
| Papillary thyroid cancer | Familial pulmonary arterial hypertension | - | | |
| Papillary thyroid cancer | Autism | True | (Liu et al., 2022; Yehia et al., 2022) | (111, 112) |
| Liver cancer | Post-traumatic stress disorder | True | phi-correlation | (1) |
| Kaposi's sarcoma | Friedreich's ataxia | True | phi-correlation | (1) |
| Prostate cancer | Glioblastoma | - | | |
| Autism | Rheumatoid arthritis | - | | |
| Borrelia burgdorferi infection | Kaposi's sarcoma | - | | |
| Multiple sclerosis | Familial dysautonomia | - | | |
| Chronic lymphocytic leukemia | Papillary thyroid cancer | True | (van der Straten et al., 2023) | (103) |
| Parkinson's disease | Colorectal cancer | - | | |
| Chronic lymphocytic leukemia | Liver cancer | - | | |
| Colorectal cancer | Friedreich's ataxia | - | | |
| Ulcerative colitis | Down syndrome | True | phi-correlation | (1) |
| Eosinophilic esophagitis | Autism | True | phi-correlation | (1) |
| Coeliac disease | Kaposi's sarcoma | True | phi-correlation | (1) |
| Cardiomyopathy | Borrelia burgdorferi infection | True | (Kuchynka et al., 2015) | (113) |
| Down syndrome | Papillary thyroid cancer | True | phi-correlation | (1) |
| Down syndrome | Cardiomyopathy | True | (Dimopoulos et al., 2023) | (114) |
| Ulcerative colitis | Liver cancer | True | phi-correlation | (1) |
| Smoker | Crohn's disease | True | phi-correlation | (1) |
| Chronic obstructive pulmonary disease | Human immunodeficiency virus infection | True | phi-correlation | (1) |
| Sessile serrated polyps | Borrelia burgdorferi infection | True | phi-correlation | (1) |
| Keratoconus | Familial dysautonomia | - | | |
| Sessile serrated polyps | Chronic lymphocytic leukemia | - | | |
| Friedreich's ataxia | Human immunodeficiency virus infection | True | phi-correlation | (1) |
| Kaposi's sarcoma | Smoker | - | | |
| Parkinson's disease | Lung cancer | - | | |
| Liver cancer | Myotonic dystrophy | True | phi-correlation | (1) |
| Sessile serrated polyps | Keratoconus | - | | |

| | | | | |
|---|---|---|---|---|
| Chronic lymphocytic leukemia | Friedreich's ataxia | - | | |
| Asthma | Liver cancer | - | | |
| Prostate cancer | Ulcerative colitis | True | (Kaneko et al., 2024; Meyers et al., 2020) | (115, 116) |
| Borrelia burgdorferi infection | Autism | - | | |
| Colorectal cancer | Asthma | True | (Guo et al., 2023) | (117) |
| Idiopathic pulmonary fibrosis | Colorectal cancer | True | (Lee et al., 2021, 2020; Stefania et al., 2023) | (72, 118, 119) |
| Schizophrenia | Idiopathic pulmonary fibrosis | - | | |
| Lung cancer | Multiple sclerosis | True | (Ge et al., 2021) | (120) |
| Liver cancer | Keratoconus | - | | |
| Friedreich's ataxia | Schizophrenia | True | phi-correlation | (1) |
| Prostate cancer | Huntington's disease | - | | |
| Ulcer | Alagille syndrome | True | phi-correlation | (1) |
| Prostate cancer | Borrelia burgdorferi infection | - | | |
| Ulcerative colitis | Familial pulmonary arterial hypertension | True | phi-correlation | (1) |
| Parkinson's disease | Crohn's disease | True | (Brudek, 2019; Kang et al., 2023) | (121, 122) |
| Cardiomyopathy | Liver cancer | - | | |
| Breast cancer | Keratoconus | - | | |
| Autism | Crohn's disease | True | (Kim et al., 2022) | (123) |
| Glioblastoma | Rheumatoid arthritis | - | | |
| Familial pulmonary arterial hypertension | Crohn's disease | - | | |
| Breast cancer | Human immunodeficiency virus infection | True | (Marino et al., 2024) | (124) |

**Table S17. Literature validation of neoplasm–neoplasm interactions in the Disease Similarity Network (DSN).** Table showing the literature validation of neoplasm–neoplasm interactions initially labeled as false positives in the DSN, following comparison with the epidemiological network from Hidalgo et al. (1) based on relative risks (RR) (Methods). It lists the disease pairs corresponding to unique ICD9 interactions. The column (In epidemiology) indicates whether the interaction was supported by epidemiological evidence, including the phi-correlation network from Hidalgo et al. or recent large-scale studies. The table also includes the source of supporting evidence (References) and the corresponding citation numbers (Ref. IDs).

| Disease 1 | Disease 2 | In epidemiology | References | Ref. IDs |
|---|---|---|---|---|
| Lung cancer | Breast cancer | True | (Curtis RE et al., 2006) | (7) |
| Adenomatous polyps | Prostate cancer | True | (Ko et al., 2016) | (63) |
| Breast cancer | Adenomatous polyps | True | (Abu-Sbeih et al., 2019) | (71) |
| Papillary thyroid cancer | Sessile serrated polyps | - | | |
| Kaposi's sarcoma | Papillary thyroid cancer | True | phi-correlation | (1) |
| Liver cancer | Breast cancer | - | | |
| Breast cancer | Papillary thyroid cancer | True | (Curtis RE et al., 2006; Trinh et al., 2021) | (7, 85) |
| Kaposi's sarcoma | Glioblastoma | True | phi-correlation | (1) |
| Papillary thyroid cancer | Lung cancer | True | phi-correlation | (1) |
| Papillary thyroid cancer | Liver cancer | True | phi-correlation | (1) |
| Colorectal cancer | Prostate cancer | True | (Curtis RE et al., 2006) | (7) |
| Kaposi's sarcoma | Breast cancer | True | (Curtis RE et al., 2006) | (7) |
| Lung cancer | Prostate cancer | - | | |
| Kaposi's sarcoma | Liver cancer | True | phi-correlation | (1) |
| Papillary thyroid cancer | Prostate cancer | True | (Tomaszewski et al., 2015) | (125) |
| Glioblastoma | Papillary thyroid cancer | True | phi-correlation | (1) |
| Liver cancer | Prostate cancer | - | | |
| Lung cancer | Colorectal cancer | True | (Airtum, 2013) | (126) |
| Adenomatous polyps | Glioblastoma | True | (Baughman et al., 1969; Paraf et al., 1997; Todd et al., 1981) | (127–129) |
| Glioblastoma | Breast cancer | True | (Raufi et al., 2017; Wei et al., 2014) | (130, 131) |
| Prostate cancer | Kaposi's sarcoma | True | phi-correlation | (1) |
| Kaposi's sarcoma | Colorectal cancer | True | phi-correlation | (1) |
| Chronic lymphocytic leukemia | Breast cancer | - | | |
| Prostate cancer | Breast cancer | True | (Barber et al., 2018; Beebe-Dimmer et al., 2015; Karlsson et al., 2006) | (132–134) |
| Lung cancer | Adenomatous polyps | - | | |
| Glioblastoma | Colorectal cancer | - | | |
| Colorectal cancer | Chronic lymphocytic leukemia | True | (Dennis and Alberts, 2007; van der Straten et al., 2023) | (102, 103) |
| Colorectal cancer | Breast cancer | True | (Curtis RE et al., 2006) | (7) |
| Papillary thyroid cancer | Colorectal cancer | True | (Sandeep et al., 2006) | (110) |
| Prostate cancer | Glioblastoma | - | | |
| Chronic lymphocytic leukemia | Papillary thyroid cancer | True | (van der Straten et al., 2023) | (103) |
| Chronic lymphocytic leukemia | Liver cancer | - | | |
| Sessile serrated polyps | Chronic lymphocytic leukemia | - | | |
